# Forecasting Alzheimer’s disease progression using non-imaging clinical features and XGBoost

**DOI:** 10.64898/2026.10.05.26364229

**Authors:** Andrew Gautier, Lingjian Zheng, Bernard G. Schreurs

## Abstract

Identifying progressive dementia during its prodromal stage is a challenging yet critical task for healthcare providers. Traditional clinical assessments are often not sensitive enough to detect warning signs of cognitive decline. Machine learning (ML) methods have shown promise in modeling clinical trajectories associated with neurodegenerative disorders such as Alzheimer’s disease (AD). Validation of these models’ forecasting abilities, however, is widely lacking in the literature. To address this gap, we established a longitudinal cohort from the National Alzheimer’s Coordinating Center (*n* = 15,727) comprising individuals who were cognitively normal (CN) or had mild cognitive impairment (MCI) at baseline. Cognitive status and clinical observations collected over up to 10 visits were used to train several ML classifiers. We employed the gradient-boosted tree ensemble method XGBoost with a custom feature-engineering pipeline. On non-imputed data, our models achieved an AUC-ROC of 0.9447 [0.9417–0.9476] and PR-AUC of 0.8554 [0.8497–0.8611] for the CN→MCI task, and an AUC-ROC of 0.9059 [0.8983–0.9134] and PR-AUC of 0.9214 [0.9156–0.9272] for the MCI→AD task. In a lead-time forecasting analysis spanning mean follow-up periods of 56.1 months (range, 9.1–214.1) for CN→MCI and 42.7 months (range, 9.1–126.1) for MCI→AD, the non-imputed models detected future progression before diagnosis in 29.58% [28.29%–30.87%] of CN→MCI progressors and 22.41% [21.36%–23.46%] of MCI→AD progressors. Among these individuals, the mean lead time was 33.04 months [32.52–33.56] and 17.95 months [17.71–18.19], respectively. False alarm rates among non-progressors were 12.6% [11.5%–13.7%] and 20.0% [18.4%–21.6%], respectively. These findings suggest that longitudinal machine learning models can identify future cognitive decline before diagnosis, indicating clinical utility for early dementia risk stratification.

**Author summary:** Alzheimer’s disease (AD) is a progressive and currently incurable neurodegenerative disorder that remains highly difficult to diagnose during its early stages. While modern early-stage dementia research relies on advanced neuroimaging, the high cost of these tests excludes many vulnerable communities. We designed our study to advance scalable, low-cost diagnostic alternatives that can identify individuals at significant risk of cognitive decline, ensuring faster access to emerging therapies.

We examined trajectories of cognitive decline in 15,727 adults (mean age = 71.45 years, SD = 9.06) who had between 2 and 10 follow-up visits. Using 37 demographic, behavioral, clinical, and genetic variables, we modeled cognitively stable and declining trajectories using the XGBoost algorithm. Additionally, we evaluated the models’ ability to forecast future diagnoses and the impact of longitudinal imputation.

Our models performed well in both classification tasks (AUC-ROC ≥ 0.90).

Imputation did not substantively alter the classifiers’ performance. In forecasting diagnosis conversion, the models raised alarms before the recorded conversion visit in a subset of progressors, with non-imputed models yielding longer net lead times than imputed models in AD forecasting. This work describes a strategy for handling mixed-length temporal dimensions in longitudinal cohorts and evaluates its prognostic ability.

## Introduction

According to the United Nations, the number of people aged 65 years or older worldwide is expected to double by 2050 [1]. This demographic shift will place an increasing burden on public health systems through a rise in age-related disorders. Alzheimer’s disease (AD), the most common cause of dementia, affects roughly 7.4 million Americans aged 65 and older in 2026 [2]. Although treatments exist to help preserve memory and motor function, there is currently no cure, and by the time AD is clinically diagnosable, the disease process is widely considered to be irreversible [2, 3].

Despite significant advancements in AD research, early and accurate diagnosis remains an enduring clinical challenge. Historically, cognitive decline was not recognized as AD until alternative causes had been ruled out, characterized as a “disease of exclusion” [4, 5]. We now understand that pathological brain changes associated with AD can precede clinical diagnosis by as much as 20 years [6]. Mild cognitive impairment (MCI) often precedes AD and is therefore a key target for predictive modeling, although many individuals with MCI do not have progressive dementia [7]. Accurate diagnosis is further complicated not only by overlapping age-related neurodegeneration and mixed dementia pathologies, but also by the inherent variance of human clinical assessment. Although standard neuropsychological assessments such as the Mini-Mental State Exam (MMSE) [8] are widely used, they have limited sensitivity for detecting subtle cognitive deficits associated with early-stage impairment [9]. In vivo imaging such as MRI or PET offers greater clinical insight, but its widespread adoption is severely hindered by high costs and lack of availability [10].

As a result, an emerging focus in AD research is on modeling the extended preclinical phase using comprehensive, low-cost datasets. Longitudinal cohorts such as the National Alzheimer’s Coordinating Center (NACC) collect repeated demographic, cognitive, and clinical assessments over time, providing longitudinal information for studying Alzheimer’s disease progression [11]. These data present several analytical challenges: repeated measurements within individuals violate the assumption of independence, missing visits and participant dropouts are common, and disease trajectories can vary substantially across individuals. Machine learning (ML) approaches capable of identifying presymptomatic patterns from routinely collected clinical data have emerged as a promising strategy for predicting disease progression, improving risk stratification, and facilitating earlier intervention [12–23]. Representative examples of ML studies are summarized in Table 1. All included studies avoid direct image-based deep learning (e.g., convolutional neural networks applied to raw MRI or PET), although several incorporate imaging-derived numerical features such as volumetric measures, cortical thickness, or PET standardized uptake value ratios.

**Table 1.** Machine-learning studies using non-imaging clinical and tabular data to classify or predict Alzheimer’s disease progression. A summary of studies applying machine learning models to structured, non-imaging data, including demographic, cognitive, clinical, and quantitative imaging-derived variables, for diagnostic classification or prediction of clinical progression across major Alzheimer’s disease cohorts. The *temporal dimension* distinguishes cross-sectional models, which use data from a single visit per participant, from longitudinal models, which incorporate repeated measurements collected over time. Abbreviations: AD = Alzheimer’s disease; MCI = mild cognitive impairment; aMCI = amnestic MCI; CN = cognitively normal; EMCI/LMCI/SMCI = early / late / stable MCI; ADNI = Alzheimer’s Disease Neuroimaging Initiative; NACC = National Alzheimer’s Coordinating Center; OASIS = Open Access Series of Imaging Studies; TADPOLE = Alzheimer’s Disease Prediction Of Longitudinal Evolution challenge dataset; ERAD = Elderly Relatives of Alzheimer’s Disease: Influence of Subjective Cognitive Decline; MADRID+90 = Factors Associated with Healthy and Pathological Aging in a Sample of Elderly People Over 90 Years Old in the City of Madrid; MLP = Multi-Layer Perceptron; LR = Logistic Regression; RF = Random Forest; KNN = k-Nearest Neighbors; NB/GaussianNB = Näıve Bayes / Gaussian Näıve Bayes; SVM = Support Vector Machine; XGBoost = Extreme Gradient Boosting; LSTM = Long Short-Term Memory network; DT = Decision Tree; LA = Linear Attention mechanism; VC = Voting Classifier; GRU = Gated Recurrent Unit; GB = Gradient Boosting; AdaBoost = Adaptive Boosting. Values reported for AUC-ROC and F1 correspond to the best-performing model described in each study. Parenthetical annotations following performance metrics identify the specific model that achieved the reported result. A dash (—) indicates that the corresponding metric was not reported by the study.

| Study | Dataset(s) | Task(s) | Temporal dimension | ML model(s) | AUC ROC | F1 score |
| --- | --- | --- | --- | --- | --- | --- |
| Adelson et al. (2023) | ADNI [N=493] | MCI→AD prediction (12-48 mo, every 6 mo) | Cross-sectional | XGBoost, KNN, MLP, LR | $\geq 0.857$ (XGBoost) | -- |
| Alatrany et al. (2024) | NACC [N not reported] | CN/MCI/AD & 4-yr follow-up | Cross-sectional | RF, KNN, NB, SVM | -- | Classification 0.854 (RF), Prediction 0.726 (RF) |
| Bogdanovic et al. (2022) | TADPOLE (from ADNI) [N=9,592] | CN/EMCI/LMCI/SMCI/AD | Cross-sectional | XGBoost, RF | -- | 0.84 (XGBoost) |
| Ding et al. (2023) | ADNI [N=347] | MCI→AD (2-yr) | Longitudinal, Cross-sectional | LSTM, RF | 0.93 (LSTM) | -- |
| Fernández-Blázquez et al. (2025) | ERAD and MADRID+90 [N=845] | CN→MCI (3-yr) | Cross-sectional | XGBoost | 0.8359 | 0.9222 |
| Kavitha et al. (2022) | OASIS [N=150] | Demented vs. non-demented | Cross-sectional | DT, SVM, RF, XGBoost, VC | -- | 0.85 (XGBoost, VC) |
| Liu et al. (2025) | NACC [N=25,179] | CN/aMCI/AD (3–10 yr) | Longitudinal | LSTM + LA., LSTM + transformer | 0.8601 (LSTM + LA.) | -- |
| Pang et al. (2023) | NACC [N>882] | CN→MCI (4-yr), MCI→AD (2, 3 yr) | Cross-sectional | SVM, RF, LR | MCI 0.714 (LR), AD $\geq 0.805$ (RF) | -- |
| Tabarestani et al. (2019) | ADNI [N=1458] | CN/MCI/AD (12, 24, 36 mo) | Longitudinal | LSTM, GRU | -- | $\geq 0.86$ (LSTM) |
| Uddin et al. (2023) | OASIS [N=150] | AD vs non-AD | Cross-sectional | GaussianNB, DT, RF, XGBoost, VC, GB | 0.96 (XGBoost) | 0.97 (XGBoost) |
| Wang et al. (2025) | NACC [N=2,379] | AD vs non-AD | Cross-sectional | RF, XGBoost, LR | 0.915 (XGBoost) | 0.833 (XGBoost) |
| Yi et al. (2023) | ADNI [N=547] | CN/MCI/AD | Cross-sectional | XGBoost, RF, Bagging, AdaBoost, NB | 0.906 (XGBoost) | -- |

XGBoost is a tree-based algorithm that has emerged as a frontrunner among these studies [24]. Adelson et al. used XGBoost to predict progression from MCI to Alzheimer’s disease over fixed windows spanning 12 to 48 months. Their model achieved an AUC-ROC of at least 0.857 across all prediction windows, outperforming a baseline classifier using only MMSE scores [12]. Similarly, Ferńandez-Bĺazquez et al. developed XGBoost-based models using exclusively non-imaging features to predict incident MCI in 845 cognitively healthy adults aged 65–87 years over a three-year follow-up, achieving an AUC-ROC of 0.8359 [16]. Some studies have reported that XGBoost performs particularly well on non-imputed datasets, outperforming traditional machine learning algorithms in various domains [25–27]. Nevertheless, the impact of preprocessing remains an active area of investigation. For example, Hidayaturrohman and Hanada found that combining MICE imputation with one-hot encoding and standardization improved XGBoost performance in heart failure prediction tasks [28]. It remains unclear whether similar preprocessing strategies provide comparable benefits in longitudinal Alzheimer’s disease cohorts, where missing data, irregular follow-up intervals, and repeated assessments are common.

Despite the inherently longitudinal nature of major Alzheimer’s cohorts such as ADNI, OASIS, and NACC, a substantial portion of these ML models rely on cross-sectional inputs drawn from a single visit per participant. This approach overlooks the temporal dimension contained in repeated assessments, including trajectories of cognitive decline and fluctuations in clinical status. By capturing changes over time, longitudinal measures may better reflect the dynamic and heterogeneous progression of Alzheimer’s disease and improve predictive performance. Several studies that did leverage longitudinal modeling required fixed-length temporal inputs [15, 20], preventing them from using additional visits beyond the prescribed sequence length.

In this study, we pursued three objectives. First, we developed a machine learning methodology for modeling cognitive trajectories in variable-length longitudinal data and evaluated its generalizability across participants who were cognitively normal (CN) or had MCI at baseline. Second, we investigated the impact of imputation on a longitudinal cohort with a high degree of missingness. Third, we evaluated the resulting models for both current-state classification and lead-time forecasting of disease progression. We aim to provide a robust and generalizable framework for longitudinal modeling in Alzheimer’s disease research.

## Methods and materials

### Dataset

We used data from versions 1–3 of the NACC Uniform Data Set (UDS), drawing on the December 2025 data freeze, which contains participants recruited across 34 Alzheimer’s Disease Research Centers between September 2005 and 2025. Before exclusions, this dataset included 55,268 unique participants. To obtain a suitable cohort, we applied several criteria. First, we restricted the sample to adults with a complete cognitive assessment history by a physician. The diagnoses were: cognitively normal (CN), mild cognitive impairment (MCI), or Alzheimer’s disease (AD). We then divided them into two subsets according to their first-visit diagnosis: CN or MCI. Within each subset, we identified non-progressors (those who maintained their initial diagnosis throughout the observation period) and progressors (those who advanced to a more severe diagnosis). To maintain adequate sample sizes while limiting class imbalance in our binary classification tasks, we further restricted the cohort to participants with between 2 and 10 consecutive visits. Finally, we excluded reverters (those who progressed during follow-up but did not end with a progression diagnosis). This resulted in a cohort of 15,727 patients. The breakdown of participants by number of visits is shown in Table 2, and the cohort selection process is summarized in Fig 1. Demographic statistics of the final cohort are presented in Table 3.

**Fig 1.**
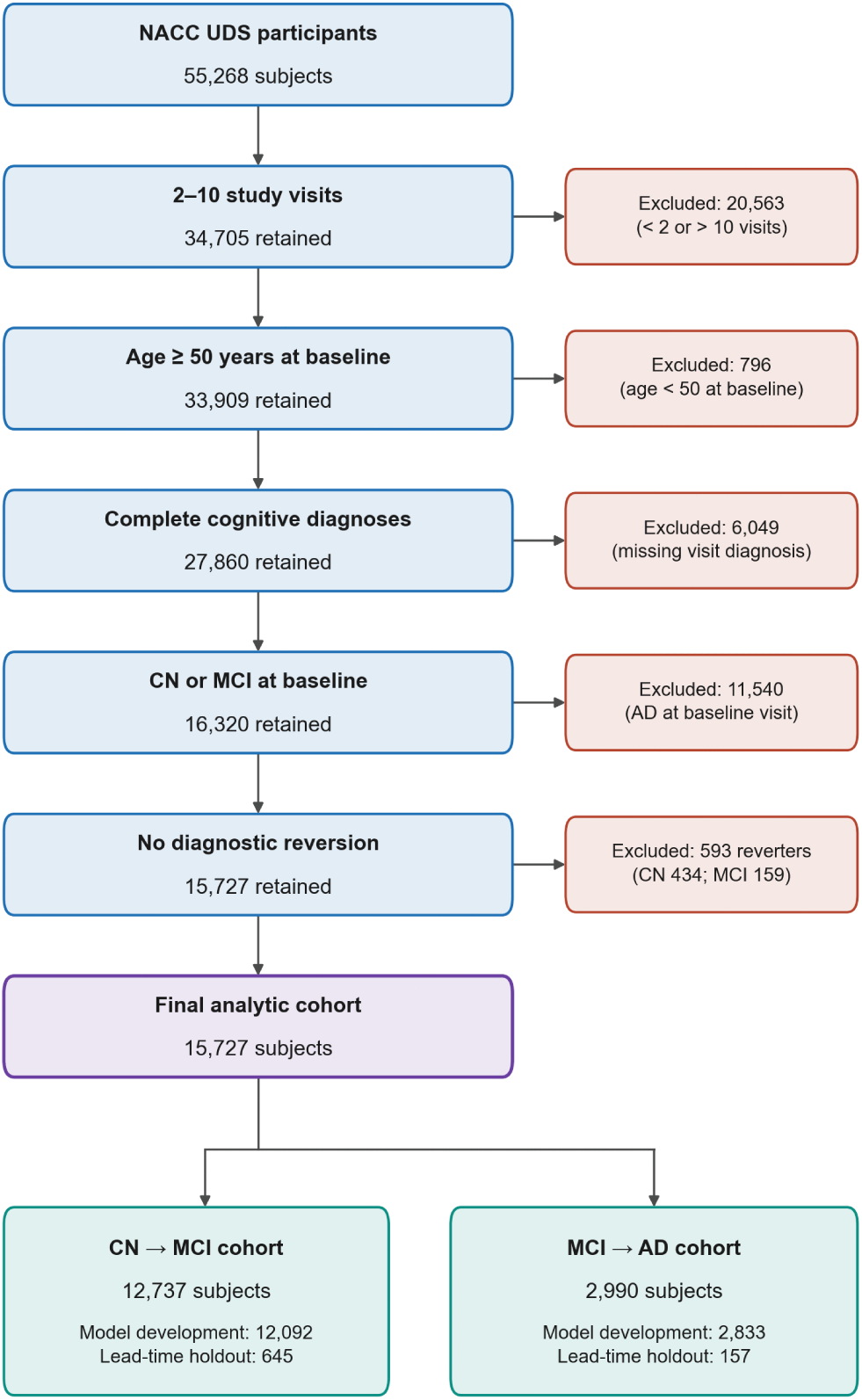
Cohort selection flowchart. Flowchart illustrating the sequential exclusion criteria applied to the initial NACC dataset. Starting from 55,268 unique patients, we retained those with 2–10 visits (34,705), with complete cognitive diagnoses (28,487), aged ≥50 years (27,880), and excluding reverters (27,287), yielding a final cohort of 15,727 participants. This cohort was then split into pooled training and 5% lead-time holdout sets.

**Table 2.**
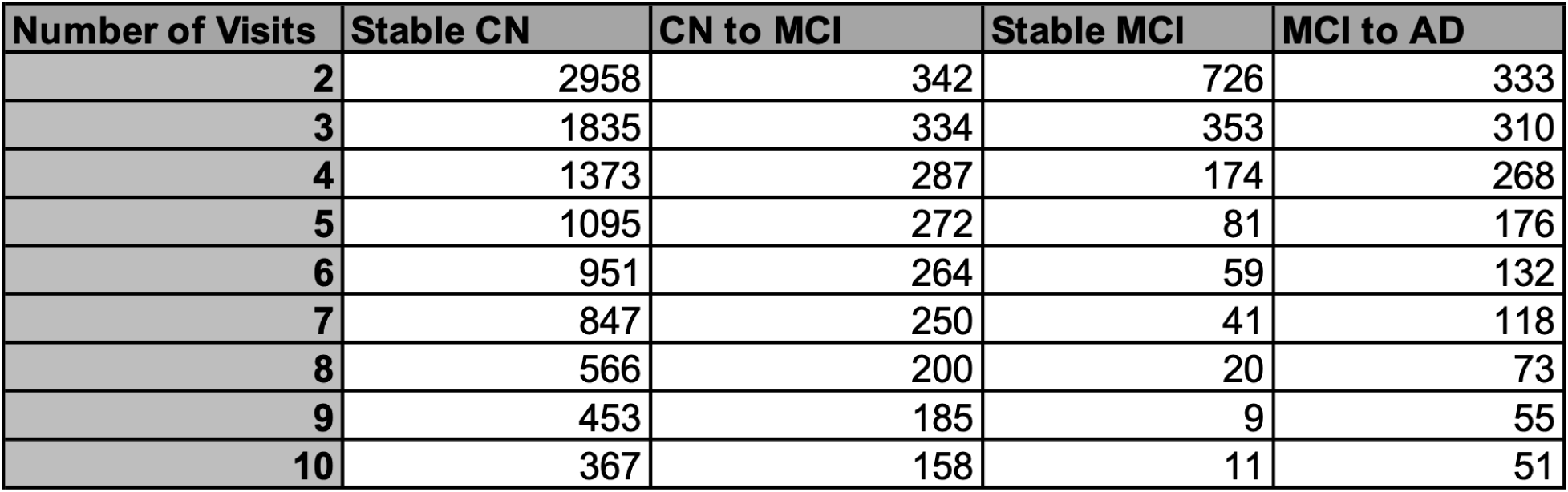
Participant counts by visit length and progression group. This figure summarizes how 15,727 participants are grouped according to their observed visit length and progression outcome, illustrating the distribution of subjects across all subgroup combinations.

| Number of Visits | Stable CN | CN to MCI | Stable MCI | MCI to AD |
| --- | --- | --- | --- | --- |
| 2 | 2958 | 342 | 726 | 333 |
| 3 | 1835 | 334 | 353 | 310 |
| 4 | 1373 | 287 | 174 | 268 |
| 5 | 1095 | 272 | 81 | 176 |
| 6 | 951 | 264 | 59 | 132 |
| 7 | 847 | 250 | 41 | 118 |
| 8 | 566 | 200 | 20 | 73 |
| 9 | 453 | 185 | 9 | 55 |
| 10 | 367 | 158 | 11 | 51 |

**Table 3.**
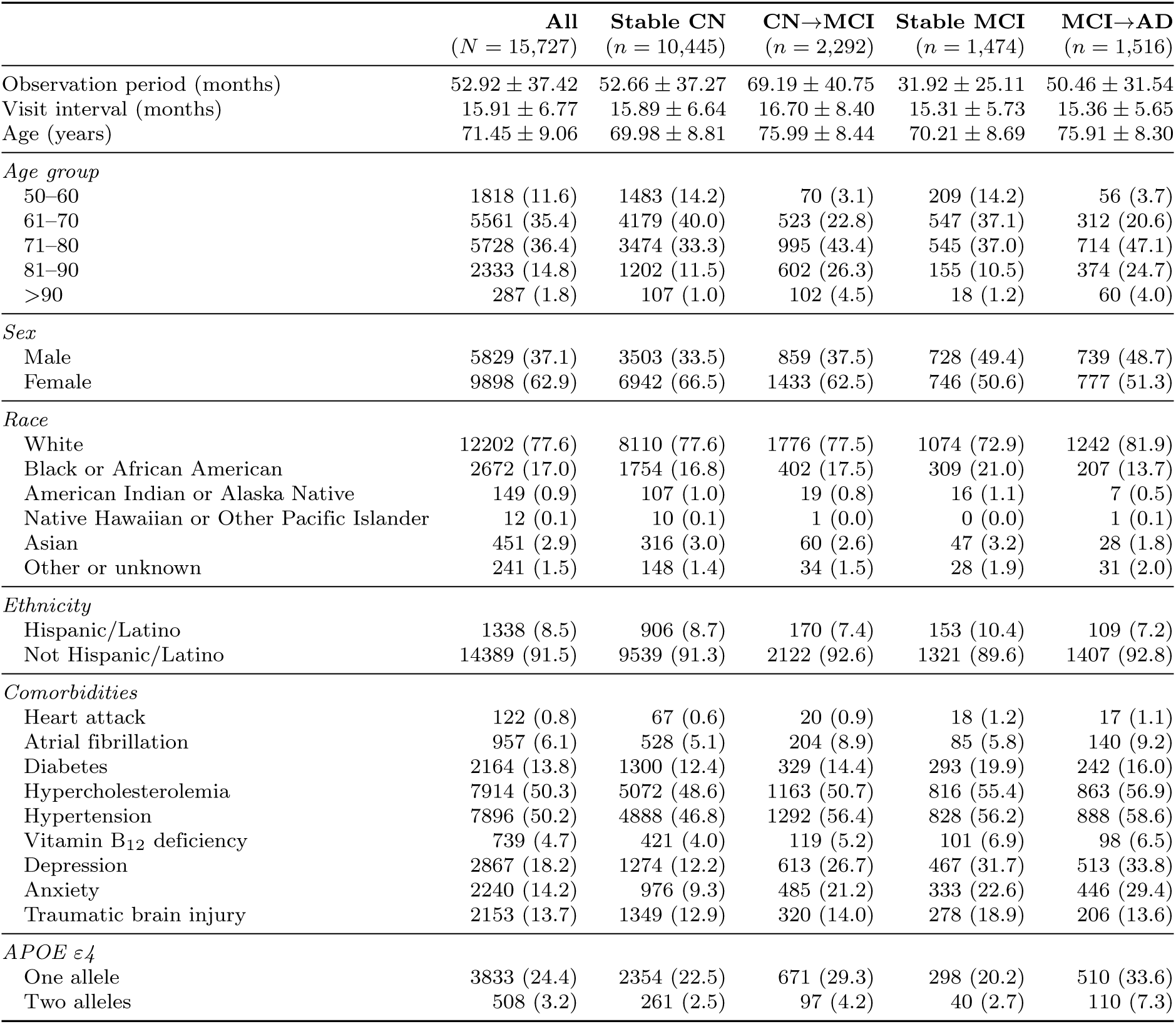
Cohort demographics for 15,727 participants, shown in summary and stratified into four progression groups (Stable CN, CN→MCI, Stable MCI, MCI→AD). Mean ± SD or *n* (%). Comorbidities are each participant’s most recently recorded status. Progressors in both cohorts were older and more often carried an APOE *ε*4 allele than non-progressors. CN→MCI progressors also more often had depression, anxiety, and hypertension, whereas comorbidity differences between stable and progressing MCI participants were mixed in direction: anxiety was more common and traumatic brain injury less common among progressors.

| | All<br>($N = 15,727$) | Stable CN<br>($n = 10,445$) | CN→MCI<br>($n = 2,292$) | Stable MCI<br>($n = 1,474$) | MCI→AD<br>($n = 1,516$) |
| --- | --- | --- | --- | --- | --- |
| Observation period (months) | 52.92 ± 37.42 | 52.66 ± 37.27 | 69.19 ± 40.75 | 31.92 ± 25.11 | 50.46 ± 31.54 |
| Visit interval (months) | 15.91 ± 6.77 | 15.89 ± 6.64 | 16.70 ± 8.40 | 15.31 ± 5.73 | 15.36 ± 5.65 |
| Age (years) | 71.45 ± 9.06 | 69.98 ± 8.81 | 75.99 ± 8.44 | 70.21 ± 8.69 | 75.91 ± 8.30 |
| *Age group* |  |  |  |  |  |
| 50–60 | 1818 (11.6) | 1483 (14.2) | 70 (3.1) | 209 (14.2) | 56 (3.7) |
| 61–70 | 5561 (35.4) | 4179 (40.0) | 523 (22.8) | 547 (37.1) | 312 (20.6) |
| 71–80 | 5728 (36.4) | 3474 (33.3) | 995 (43.4) | 545 (37.0) | 714 (47.1) |
| 81–90 | 2333 (14.8) | 1202 (11.5) | 602 (26.3) | 155 (10.5) | 374 (24.7) |
| >90 | 287 (1.8) | 107 (1.0) | 102 (4.5) | 18 (1.2) | 60 (4.0) |
| *Sex* |  |  |  |  |  |
| Male | 5829 (37.1) | 3503 (33.5) | 859 (37.5) | 728 (49.4) | 739 (48.7) |
| Female | 9898 (62.9) | 6942 (66.5) | 1433 (62.5) | 746 (50.6) | 777 (51.3) |
| *Race* |  |  |  |  |  |
| White | 12202 (77.6) | 8110 (77.6) | 1776 (77.5) | 1074 (72.9) | 1242 (81.9) |
| Black or African American | 2672 (17.0) | 1754 (16.8) | 402 (17.5) | 309 (21.0) | 207 (13.7) |
| American Indian or Alaska Native | 149 (0.9) | 107 (1.0) | 19 (0.8) | 16 (1.1) | 7 (0.5) |
| Native Hawaiian or Other Pacific Islander | 12 (0.1) | 10 (0.1) | 1 (0.0) | 0 (0.0) | 1 (0.1) |
| Asian | 451 (2.9) | 316 (3.0) | 60 (2.6) | 47 (3.2) | 28 (1.8) |
| Other or unknown | 241 (1.5) | 148 (1.4) | 34 (1.5) | 28 (1.9) | 31 (2.0) |
| *Ethnicity* |  |  |  |  |  |
| Hispanic/Latino | 1338 (8.5) | 906 (8.7) | 170 (7.4) | 153 (10.4) | 109 (7.2) |
| Not Hispanic/Latino | 14389 (91.5) | 9539 (91.3) | 2122 (92.6) | 1321 (89.6) | 1407 (92.8) |
| *Comorbidities* |  |  |  |  |  |
| Heart attack | 122 (0.8) | 67 (0.6) | 20 (0.9) | 18 (1.2) | 17 (1.1) |
| Atrial fibrillation | 957 (6.1) | 528 (5.1) | 204 (8.9) | 85 (5.8) | 140 (9.2) |
| Diabetes | 2164 (13.8) | 1300 (12.4) | 329 (14.4) | 293 (19.9) | 242 (16.0) |
| Hypercholesterolemia | 7914 (50.3) | 5072 (48.6) | 1163 (50.7) | 816 (55.4) | 863 (56.9) |
| Hypertension | 7896 (50.2) | 4888 (46.8) | 1292 (56.4) | 828 (56.2) | 888 (58.6) |
| Vitamin B<sub>12</sub> deficiency | 739 (4.7) | 421 (4.0) | 119 (5.2) | 101 (6.9) | 98 (6.5) |
| Depression | 2867 (18.2) | 1274 (12.2) | 613 (26.7) | 467 (31.7) | 513 (33.8) |
| Anxiety | 2240 (14.2) | 976 (9.3) | 485 (21.2) | 333 (22.6) | 446 (29.4) |
| Traumatic brain injury | 2153 (13.7) | 1349 (12.9) | 320 (14.0) | 278 (18.9) | 206 (13.6) |
| *APOE ε4* |  |  |  |  |  |
| One allele | 3833 (24.4) | 2354 (22.5) | 671 (29.3) | 298 (20.2) | 510 (33.6) |
| Two alleles | 508 (3.2) | 261 (2.5) | 97 (4.2) | 40 (2.7) | 110 (7.3) |

### Ethics statement

The Institutional Review Board of West Virginia University waived ethical approval for this work. The study (IRB ID STUDY00000853) was determined to be exempt under exemption category 4 (secondary research on data or specimens, no consent required) on October 5, 2026.

### Variable selection and preprocessing

Variables were selected with guidance from the 2024 Lancet Commission report on dementia risk factors [29] and clinical domain knowledge, covering comorbidities, cognitive assessments, behavioral factors, demographics, and Instrumental Activities of Daily Living (IADLs). These categories capture both static patient characteristics and longitudinal disease trajectories. A full list of the variables used can be found in S1.

### Static variables

The 17 static variables comprised eight baseline characteristics and nine comorbidities. The baseline characteristics were age at baseline (years), sex, race, Hispanic/Latino ethnicity, years of education, total years of smoking, family history of cognitive impairment in a first-degree relative, and APOE *ε*4 allele count (0, 1, or 2 copies); family history and APOE *ε*4 were included because of their well-replicated associations with Alzheimer’s disease risk [30, 31]. The comorbidities were myocardial infarction, atrial fibrillation, diabetes, hypercholesterolemia, hypertension, vitamin B12 deficiency, active depression, active anxiety, and traumatic brain injury. Each static variable took the participant’s most recent recorded value; visits at which an item was not collected, such as the health history at UDS version 3 follow-up visits, were skipped.

Baseline characteristics were used as recorded, apart from setting sentinel codes to missing; family history was coded 1 if reported and 0 if absent or unknown. Comorbidities were recoded as binary, with 1 indicating an active or recent condition and 0 an absent, remote/inactive, or unknown one.

### Longitudinal variables

Visit-level variables provided a way to model cognitive trajectories. Cognitive assessments included the Mini-Mental State Examination (MMSE) [8], the Geriatric Depression Scale (GDS) [32], and the Clinical Dementia Rating (CDR^®^) Sum of Boxes [33], a well-established instrument for staging dementia severity in clinical trials. UDS version 3 replaced the MMSE with the Montreal Cognitive Assessment (MoCA) [34]; at visits without an MMSE, the education-adjusted MoCA score was converted to the MMSE scale with the equipercentile crosswalk of the NACC Crosswalk Study [35], so that about 98% of participants had a cognitive screening score at one or more visits. Social functioning was represented by living situation (alone, with a spouse or partner, with a relative or friend, or in a group setting) and the CDR Community Affairs domain score (COMMUN). Hearing and vision impairment were included, as decline in one or more of these senses has a correlation with dementia [36]. Behavioral risk factors comprised body mass index (BMI), alcohol use, and past-30-day tobacco use. IADLs were drawn from the Functional Activities Questionnaire (FAQ), including paying bills, shopping, meal preparation, tracking events, and remembering appointments. For temporal reference, each visit included a months-since-baseline value.

The preprocessing pipeline transforms raw subject-visit records into a clean, analysis-ready dataset.

Missing values are systematically handled: sentinel codes (e.g., -4, 88, 888) are converted to NaN, and imputation can be applied at this phase as well. Hearing and vision composites are derived from six raw sensory variables using rule-based classification into ordinal scores (0 = normal, 1 = impaired with effective aid, 2 = impaired without effective aid). See Fig S1.1 for details. Subjects are filtered to include only those with between min visits (default 2) and max visits (10) visits, no “Unknown” diagnostic labels, and age at baseline ≥ 50 years.

### Ground truth labeling

For each subject, the binary target Prog ID (progression status) is set to 1 if any visit shows a more advanced cognitive state than the first visit (i.e., progression from CN to MCI/AD or from MCI to AD), and 0 otherwise. A stratified 5% lead-time holdout is randomly sampled from each Prog ID group within each visit-count stratum for the lead-time forecasting analysis. The final output includes separate pooled datasets for CN-starting and MCI-starting subjects, each containing static variables, longitudinal lists, and the derived composites.

### Missing values

We explored the degree of missingness in this new set of variables after applying our initial recoding. Because XGBoost handles missing values robustly, we did not apply any additional criteria of completeness in the records. Supplementary information on the missingness in the dataset can be found in S2.

### Feature engineering

Our challenge in feature engineering was to transform per-subject longitudinal visit records into a fixed-width, visit-count-agnostic feature vector suitable for gradient-boosted tree classification. Because patients have differing numbers of visits, all features are summary statistics that collapse the visit dimension. The model therefore generalizes across patients with 2 visits and patients with 10 visits without requiring imputation or fixed-length padding. The pipeline extracts summary statistics from these lists, yielding a single row per subject regardless of the number of visits observed. Categorical variables (sex, race, APOE *ε*4 allele count, and the first and last living-situation codes) were declared categorical to XGBoost, which splits them by groups of categories rather than by numeric order.

For each of the 19 numeric longitudinal variables, we computed nine standard summary statistics (mean, maximum, minimum, standard deviation, range, first, last, last-minus-first, and number of valid visits), together with time-normalized slope and acceleration estimated by ordinary least squares regression against months since baseline rather than visit index. This ensures that rate features are expressed in clinical units per month and remain comparable across patients with irregular visit spacing. Two additional features, the standard deviation of pairwise per-month slopes std slope and the maximum relative change between consecutive visits pct change, captured non-linear trajectories and discrete step-changes. Living situation is nominal, so instead of arithmetic summaries it contributed its first and last recorded codes and a flag for any change. Because months since baseline is itself the time axis, only summary and visit-spacing features (the mean and standard deviation of inter-visit intervals) were derived from it. Lagged features, encoding the second-, third-, and fourth-to-last valid observations, were computed for seven clinically prioritized variables—MMSE, CDR Sum of Boxes, GDS, hearing, vision, bill-paying, and shopping—to preserve recent temporal ordering that aggregate statistics otherwise discard.

Special consideration was given to sensory and behavioral variables. Dual sensory impairment, concurrent hearing and vision loss, is a well-documented risk factor for dementia, and its effect is reported to be multiplicative rather than additive [36]. A model that treats hearing and vision as independent inputs cannot capture this synergy, so three explicit interaction terms (product, sum, and mean) were computed from the engineered *last* values of each sensory composite. Alcohol use, whose NACC coding is non-monotonic (0 = absent, 1 = recent/active, 2 = remote/inactive), was recoded so that higher values consistently indicate greater clinical concern before any summary or slope features were derived.

In total, this produced 302 features: 17 static variables, 209 longitudinal summary features (11 statistics across 19 variables), 19 slope-variability features, 19 percentage-change features, 3 living-situation features, 11 visit-timing features, 21 lagged features, and 3 sensory interaction terms. Features that require more observations than a patient has (e.g., acceleration requires ≥ 4 visits) were set to NaN rather than imputed, relying on XGBoost’s native handling of missing values. No scaling or normalization was applied, as XGBoost is scale-invariant. An exhaustive description of the feature engineering process can be found in S3.

### Training and validation

For each conversion task (CN→MCI and MCI→AD), model development was conducted across 20 independent experimental variants. In the non-imputed analyses, 10 stratified 80/20 train–test splits were generated using 10 different random seeds drawn from a fixed master seed. For the imputed analyses, a further 10 stratified 80/20 splits were generated in the same manner (seeds 42–51), and Multiple Imputation by Chained Equations (MICE) was fit separately for each variant on the training cohort only and then applied to both the training and test cohorts, preventing information leakage. Within each variant, feature engineering was performed independently on the training and test partitions, so that no statistics derived from the test set influenced model development.

For each variant, a gradient-boosted decision tree classifier (XGBoost) was trained, with class imbalance addressed by weighting the positive class by the negative-to-positive ratio. Hyperparameters were optimized using Bayesian optimization implemented with Optuna and a Tree-structured Parzen Estimator (TPE) sampler. The search space included the number of boosting rounds, maximum tree depth, learning rate, row and column subsampling fractions, minimum child weight, gamma, and L1/L2 regularization parameters. Candidate configurations were evaluated using five-fold stratified cross-validation conducted exclusively within the training cohort, with mean area under the receiver operating characteristic curve (AUC-ROC) as the optimization objective. For each dataset variant, 1,000 optimization trials were performed, and the highest-performing configuration was selected for final model development.

Following hyperparameter optimization, a final XGBoost classifier was trained for each variant using the optimal configuration identified within that variant, yielding 10 final models for the non-imputed analyses and 10 for the imputed analyses for each conversion task. Each model was evaluated on its corresponding held-out test cohort, and performance metrics were calculated independently and then aggregated across variants to quantify variability attributable to train–test partitioning and multiple imputation. Confidence intervals for all reported performance metrics were estimated from the between-run sample standard deviation using the *t*-distribution.

### Prognostic ability

To evaluate the temporal relationship between model predictions and subsequent clinical progression, we performed a lead-time analysis on the stratified 5% lead-time holdout described above. Separate analyses were conducted for the CN→MCI and MCI→AD tasks using the non-imputed and MICE-imputed pipelines and their respective trained models.

For each subject, the visit history was decomposed into a series of successively longer prefixes, from the first two visits through the complete record, and the classifiers trained in the primary analysis were applied to every prefix. Predictions were therefore generated at every available follow-up, beginning with the second visit (mean 15.6 months after baseline in the CN→MCI cohort and 15.1 months in the MCI→AD cohort) and extending through the final visit for progressors or the last unmasked visit for controls. Only the longitudinal variables were truncated: every prefix carried the participant’s static values, which for time-varying items (comorbidities, years of smoking, and years of education) are the most recent recorded values and may postdate the prefix. These items accounted for less than 2% of the SHAP attribution in either task. Across the lead-time holdout, follow-up spanned a mean of 56.1 months (range, 9.1–214.1 months) for the CN→MCI experiment and 42.7 months (range, 9.1–126.1 months) for the MCI→AD experiment; these windows were identical for the non-imputed and MICE-imputed pipelines, which shared the same visit schedules. For progressors, lead time was defined as the number of months between the earliest prefix for which the predicted probability exceeded the decision threshold and the first visit at which clinical conversion was recorded, such that positive values indicate alarms preceding conversion. For non-progressors, any alarm on a truncated history constituted a false positive, and the false-alarm rate was defined as the fraction of non-progressor subjects with at least one alarm. Sensitivity was the fraction of progressors with an alarm on any prefix. For the subject-level AUC-ROC and PR-AUC (Table 5), each participant was scored by their highest predicted probability over the evaluated prefixes; for progressors these include prefixes ending at or after conversion, so these areas measure discrimination over the whole record rather than ahead of conversion.

Decision thresholds were selected using sensitivity, specificity, and false-alarm-rate profiles. Thresholds of 0.25 for the CN→MCI model and 0.30 for the MCI→AD model provided favorable trade-offs between early detection performance and false-positive rate and were used for all subsequent lead-time analyses.

Because non-progressors whose records end shortly before an unobserved conversion could otherwise be misclassified as stable, the final visits of each non-progressor history were masked, i.e., excluded from evaluation. For each candidate threshold, the mask length was derived from the progressor lead-time distribution. The 80th percentile of early lead times was divided by the cohort’s mean inter-visit interval (approximately 15.9 months; the second visit occurred a mean of 15.6 and 15.1 months after baseline in the CN→MCI and MCI→AD holdouts, respectively) and rounded to the nearest visit. At the selected thresholds, this procedure yielded mask lengths of three visit intervals for CN→MCI and two visit intervals for MCI→AD, balancing the exclusion of near-future converters against retention of an adequate control sample. Additional justification for the threshold and mask-length selection, together with a detailed description of cohort construction, sequence truncation, and lead-time computation, is provided in S4.

### Computational environment

All experiments were conducted on a MacBook Pro with an Apple M4 chip and 24 GB of RAM using Python 3.14.4. Models were trained with XGBoost 3.2.0 [24], with training parallelized across CPU cores using joblib 1.5.3 [37]. Data preprocessing, model evaluation, and hyperparameter tuning used scikit-learn 1.8.0 [38], and hyperparameter optimization used Optuna 4.9.0 [39]. Data manipulation and statistical analysis relied on pandas 3.0.3 [40], NumPy 2.4.6 [41], and SciPy 1.17.1 [42]. SHAP values were computed with SHAP 0.52.0 [43], and figures were produced with Matplotlib 3.10.9 [44] and seaborn 0.13.2 [45].

## Results

### Classifiers

Table 4 reports classification performance for the CN→MCI and MCI→AD tasks under both pipelines. Across the *n* = 10 repeated runs, each metric is summarized as the mean *x̄* with a 95% confidence interval 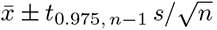, where *s* is the between-run sample standard deviation.

**Table 4.** Full classification performance of XGBoost models. Evaluation metrics for the CN MCI and MCI AD conversion tasks under non-imputed and MICE-imputed pipelines. Reported metrics include area under the receiver operating characteristic curve (AUC-ROC), area under the precision-recall curve (PR-AUC), accuracy, sensitivity (recall), specificity, positive predictive value (PPV), negative predictive value (NPV), and F1 score. Threshold-based metrics use a predicted probability of 0.5. Values are the mean across independent runs with 95% confidence intervals. CN = cognitively normal; MCI = mild cognitive impairment; AD = Alzheimer’s disease; MICE = Multiple Imputation by Chained Equations.

| Imputed | CN→MCI |  | MCI→AD |  |
| --- | --- | --- | --- | --- |
|  | No | Yes | No | Yes |
| AUC-ROC | 0.945 (0.942–0.948) | 0.946 (0.941–0.951) | 0.906 (0.898–0.913) | 0.902 (0.895–0.909) |
| PR-AUC | 0.855 (0.850–0.861) | 0.856 (0.847–0.866) | 0.921 (0.916–0.927) | 0.918 (0.913–0.924) |
| Sensitivity | 0.860 (0.849–0.870) | 0.869 (0.856–0.883) | 0.800 (0.784–0.816) | 0.765 (0.746–0.783) |
| Specificity | 0.913 (0.903–0.923) | 0.911 (0.904–0.918) | 0.850 (0.836–0.864) | 0.855 (0.842–0.868) |
| PPV | 0.686 (0.662–0.711) | 0.683 (0.668–0.699) | 0.847 (0.835–0.859) | 0.845 (0.834–0.856) |
| NPV | 0.967 (0.965–0.969) | 0.970 (0.967–0.972) | 0.805 (0.792–0.818) | 0.779 (0.766–0.792) |
| F1 | 0.762 (0.752–0.773) | 0.765 (0.756–0.774) | 0.822 (0.812–0.833) | 0.802 (0.792–0.813) |
| Accuracy | 0.903 (0.897–0.910) | 0.904 (0.899–0.909) | 0.825 (0.815–0.835) | 0.809 (0.800–0.818) |

**Table 5.** Full lead-time forecasting performance of XGBoost models. Lead-time analysis metrics for the CN MCI and MCI AD conversion tasks under non-imputed and MICE-imputed pipelines, evaluated at decision thresholds of 0.25 (CN MCI) and 0.30 (MCI AD) with control mask lengths of three and two visit intervals, respectively. Metrics include AUC-ROC, PR-AUC, sensitivity, specificity, positive predictive value (PPV), negative predictive value (NPV), accuracy, F1 score, false alarm rate (FAR), mean lead time, and net lead time in months. Values are the mean across runs with 95% confidence intervals.

| Imputed | CN→MCI |  | MCI→AD |  |
| --- | --- | --- | --- | --- |
|  | No | Yes | No | Yes |
| AUC-ROC | 0.959 (0.957–0.960) | 0.962 (0.960–0.965) | 0.926 (0.920–0.932) | 0.915 (0.908–0.922) |
| PR-AUC | 0.953 (0.952–0.953) | 0.954 (0.952–0.957) | 0.978 (0.976–0.980) | 0.973 (0.971–0.975) |
| Sensitivity | 0.931 (0.921–0.941) | 0.941 (0.937–0.946) | 0.880 (0.868–0.891) | 0.888 (0.881–0.895) |
| Specificity | 0.874 (0.863–0.885) | 0.868 (0.863–0.873) | 0.800 (0.784–0.816) | 0.819 (0.800–0.838) |
| PPV | 0.802 (0.790–0.815) | 0.798 (0.792–0.804) | 0.938 (0.933–0.943) | 0.945 (0.939–0.950) |
| NPV | 0.959 (0.953–0.964) | 0.965 (0.962–0.967) | 0.661 (0.638–0.683) | 0.681 (0.664–0.699) |
| F1 | 0.862 (0.857–0.867) | 0.863 (0.859–0.867) | 0.908 (0.900–0.915) | 0.915 (0.910–0.920) |
| Accuracy | 0.894 (0.889–0.899) | 0.894 (0.891–0.897) | 0.862 (0.851–0.872) | 0.873 (0.866–0.880) |
| FAR | 0.126 (0.115–0.137) | 0.132 (0.127–0.137) | 0.200 (0.184–0.216) | 0.181 (0.162–0.200) |
| Mean lead time (months) | 33.04 (32.52–33.56) | 32.44 (31.63–33.26) | 17.95 (17.71–18.19) | 18.24 (17.85–18.62) |
| Net lead time (months) | 9.92 (9.38–10.46) | 10.02 (9.59–10.46) | 1.64 (1.10–2.18) | 0.55 (0.21–0.89) |

Using the confusion-matrix counts (true positives TP, false positives FP, false negatives FN, and true negatives TN), the threshold-based metrics were computed as

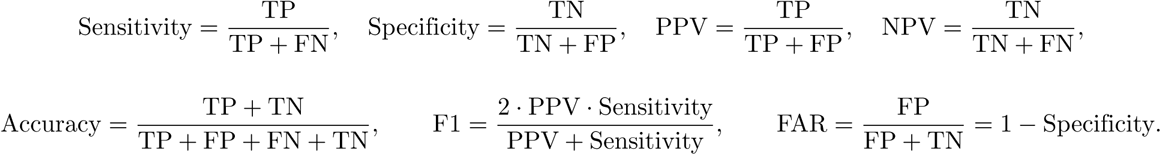

AUC-ROC and PR-AUC are the areas under the receiver operating characteristic and precision–recall curves, respectively, and are shown in Fig 2.

**Fig 2.**
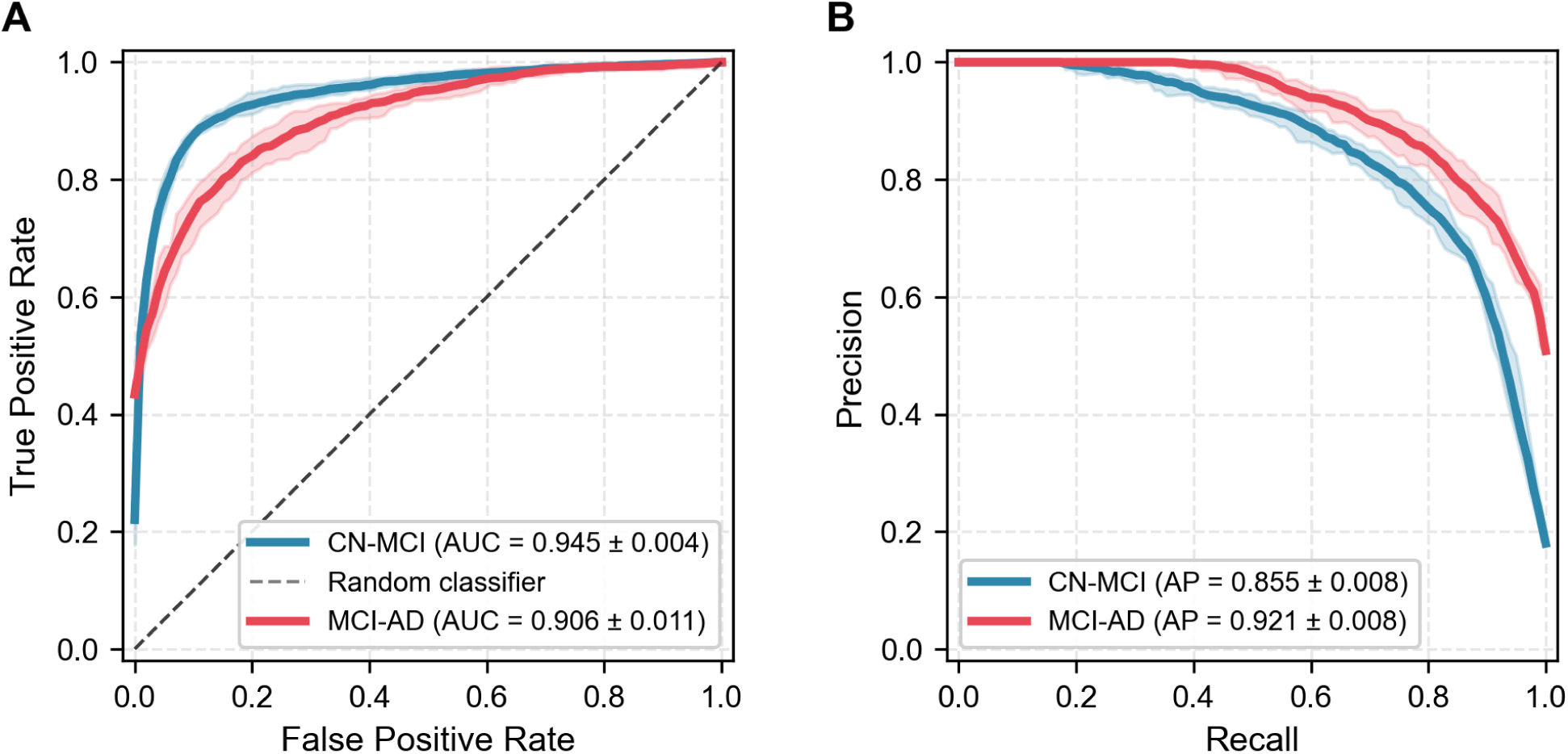
Performance of XGBoost models trained on non-imputed data for predicting cognitive conversion. (A) Receiver operating characteristic (ROC) curves for predicting conversion from cognitively normal (CN) to mild cognitive impairment (MCI; blue) and from MCI to Alzheimer’s disease (AD; red). (B) Corresponding precision-recall (PR) curves for the same two tasks. Solid lines indicate the mean performance aggregated across ten independent runs (random train-test splits), and shaded ribbons represent the 95% confidence interval estimated across these runs. In panel A, the dashed diagonal line represents the performance of a random classifier (area under the curve = 0.5). Chance-level average precision, equal to the positive-class prevalence in the test sets, was 0.180 for CN→MCI and 0.508 for MCI→AD. Legend values are the mean ± SD across runs. AUC = area under the ROC curve; AP = average precision.

### Feature importance and effect of imputation

Feature importance was computed as each variable’s share of the total gain, that is, the loss reduction from all splits on features derived from that variable, averaged across the ten non-imputed runs (Fig 3). The CDR Sum of Boxes (CDRSUM) ranked first in both tasks, with 60% of the total gain for CN→MCI and 43% for MCI→AD, followed by NACCMMSE (10% in both). For CN→MCI, NACCBMI and NACCGDS ranked third and fourth; for MCI→AD, the CDR Community Affairs score (COMMUN) ranked third and past-30-day tobacco use (TOBAC30) fifth. Among individual derived features (Fig S6.5 in S6), the most informative were CDR-SB descriptors: the most recent value (CDRSUM last) and the total change over follow-up (CDRSUM last minus first) ranked in the top three in both tasks, together with the time-normalized slope (CDRSUM slope) for CN→MCI and the maximum (CDRSUM max) for MCI→AD.

**Fig 3.**
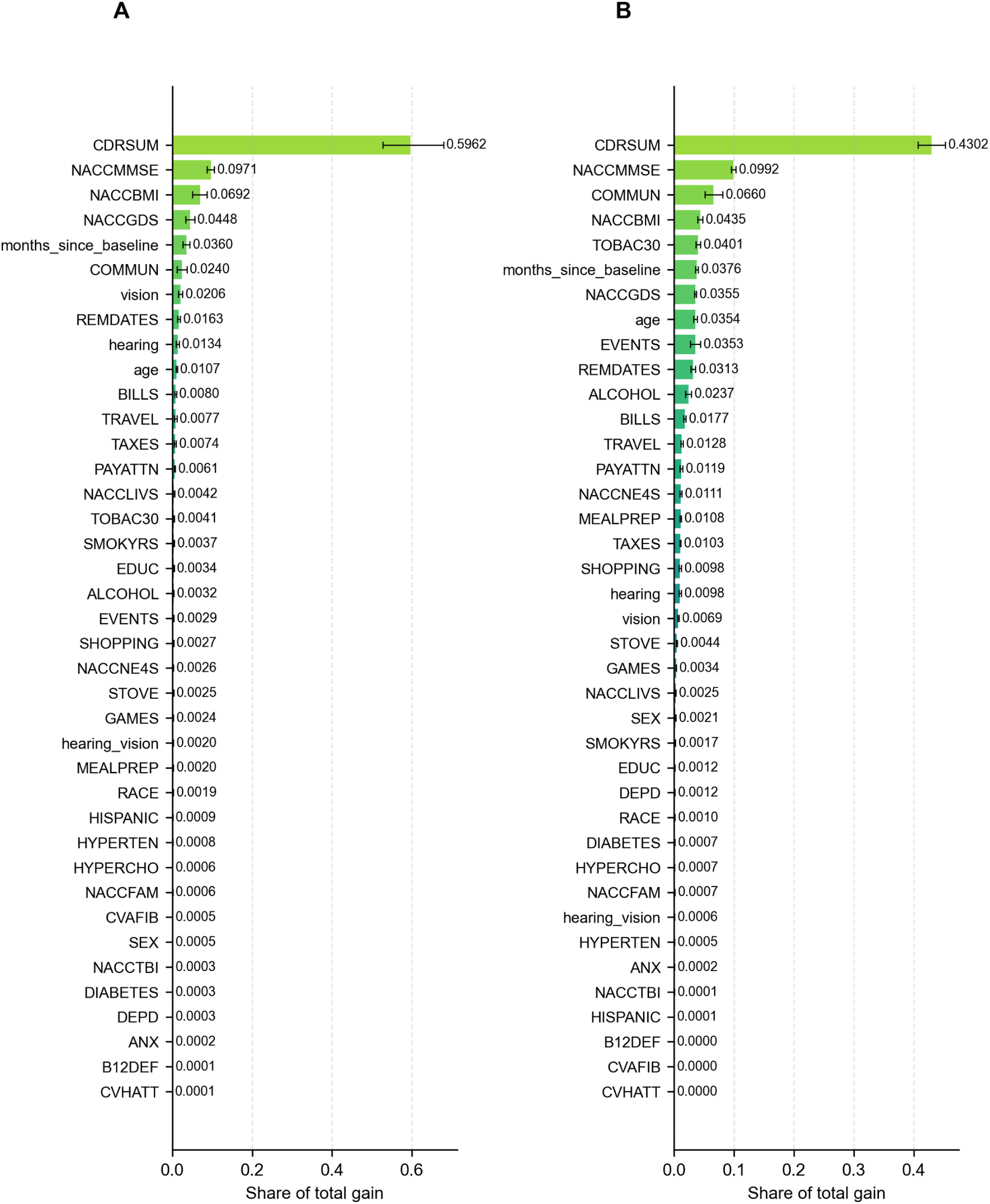
Feature importance for the longitudinal XGBoost classifiers. All 39 variables ranked by their share of the total gain, averaged across the ten non-imputed runs, for (A) CN MCI and (B) MCI AD classification. A variable’s total gain is the loss reduction from all splits on the features derived from it (e.g., CDRSUM slope and CDRSUM last). Bars show the mean share across runs and error bars denote the 95% percentile bootstrap confidence interval of the mean (2,000 resamples of the ten runs).

SHAP summary plots, with SHAP values likewise summed over each variable’s derived features (Fig 4 and Fig 5), also ranked CDRSUM and NACCMMSE first and second in both tasks, followed by NACCBMI and NACCGDS for CN→MCI and by age and TOBAC30 for MCI→AD. In each SHAP plot, dots to the right of zero pushed that prediction toward conversion and dots to the left pushed it away; a variable whose red (high-value) dots lie to the right therefore raises predicted risk when high, and a wider horizontal spread marks a larger influence on individual predictions. SHAP values are additive across a variable’s derived features and across variables, which supports the comparisons below; the feature grouping, summation, and beeswarm construction are described in S6.

**Fig 4.**
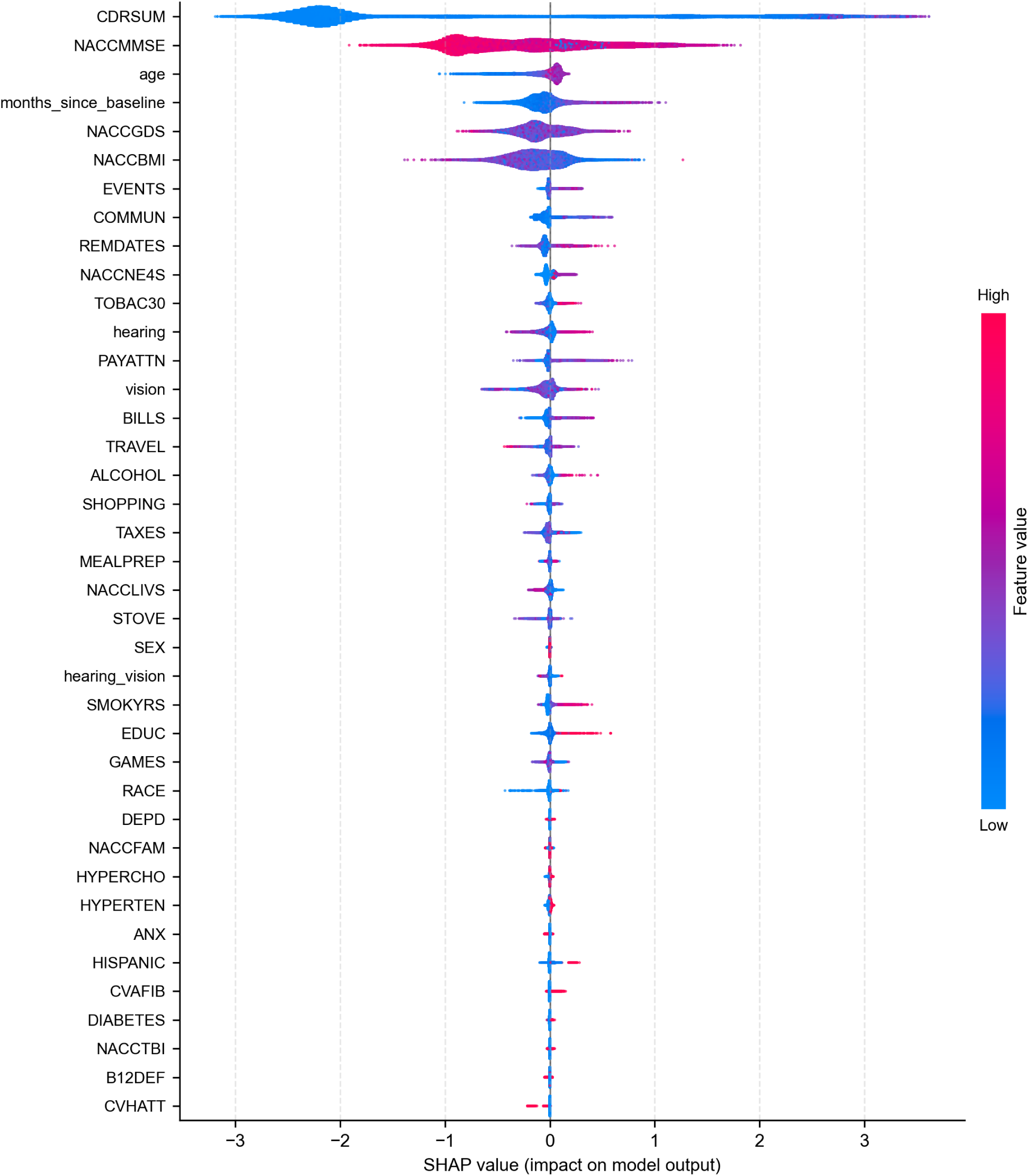
SHAP summary plot for the CN→MCI classifier. All 39 variables, with SHAP values computed on training samples pooled across the ten non-imputed runs and summed across all features derived from each variable. Each dot represents a single training prediction; horizontal position indicates the variable’s contribution to the predicted log-odds, and dot color indicates its value from low to high (for variables with several derived features, a SHAP-weighted average of their min–max normalized values). Rows are in the same order in Figs 4, 5, S6.3, and S6.4, sorted by the mean share of SHAP attribution over both tasks and pipelines, so a variable’s row need not match its rank in this task (Table S6.1).

**Fig 5.**
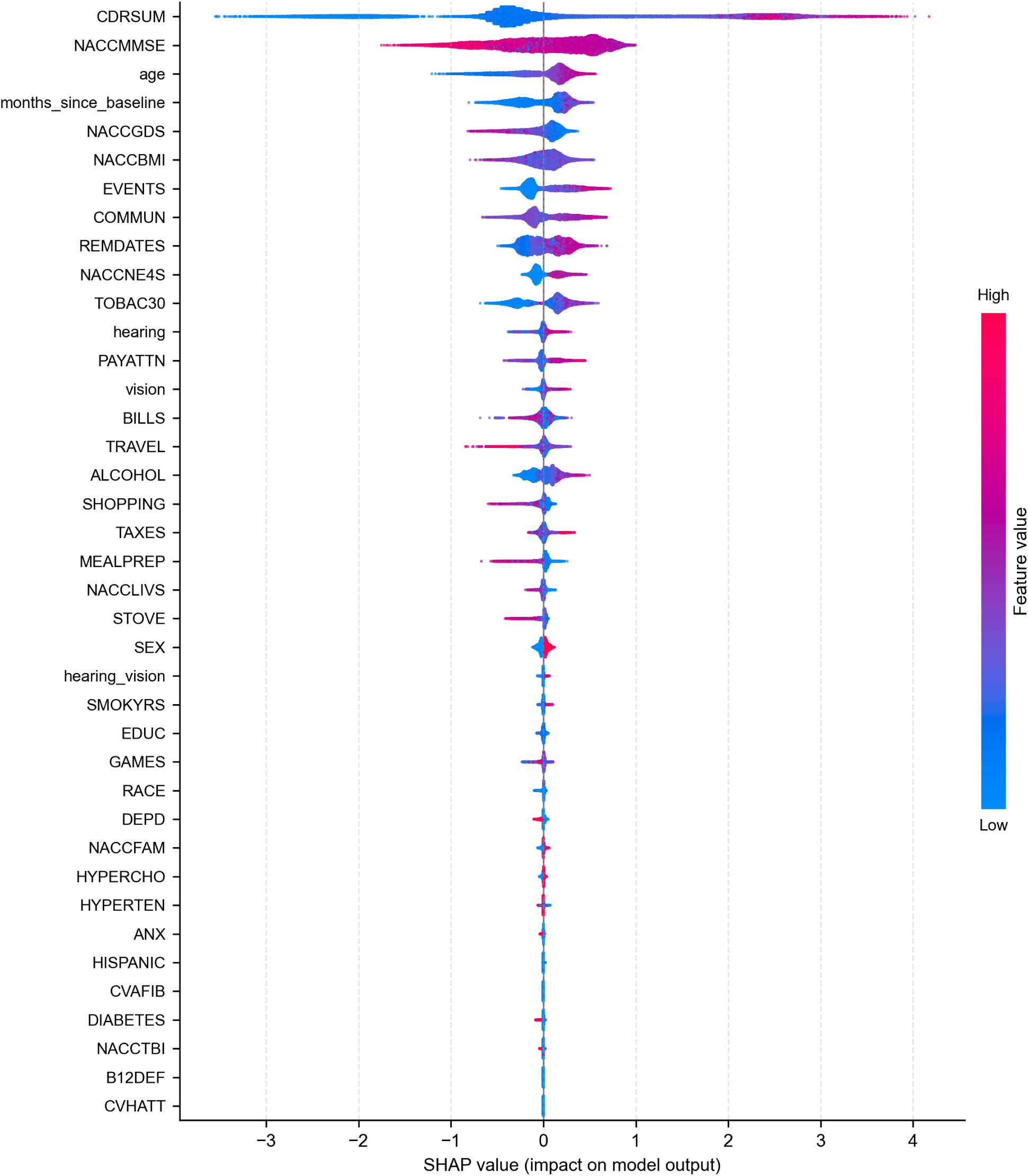
SHAP summary plot for the MCI→AD classifier. All 39 variables, with SHAP values computed on training samples pooled across the ten non-imputed runs and summed across all features derived from each variable. Each dot represents a single training prediction; horizontal position indicates the variable’s contribution to the predicted log-odds, and dot color indicates its value from low to high (for variables with several derived features, a SHAP-weighted average of their min–max normalized values). Rows are in the same order as in Fig 4.

The two tasks drew on different information (Table S6.1 in S6). CDRSUM accounted for 53% of the SHAP attribution for CN→MCI but 29% for MCI→AD, whereas the ten FAQ items and COMMUN together rose from 8% to 24%; both differences held under imputation. Across all variables, the shares of the two tasks were nonetheless correlated (Spearman *ρ* = 0.79). Higher CDRSUM, lower NACCMMSE, older age, more APOE *ε*4 alleles, and a lower most recent BMI raised predicted risk in both tasks. The effect of depressive symptoms reversed between tasks: a rising GDS score raised the predicted risk of MCI, whereas a high GDS score lowered the predicted risk of AD. By feature type, trajectory features (slopes, change since the first visit, range, variability, percentage change, and acceleration) received 44% (CN→MCI) and 45% (MCI→AD) of the attribution, compared with 46% and 38% for level features, and the change in CDRSUM since the first visit (CDRSUM last minus first) was the fourth and second most important individual feature. Lagged values (5% and 4%), acceleration (3% and 2%), and the hearing–vision interaction terms (0.2% or less) contributed little.

Imputation had little effect on classification performance. For CN→MCI, the MICE-imputed models achieved an AUC-ROC of 0.9457 [0.9408–0.9506] and a PR-AUC of 0.8563 [0.8467–0.8659], compared with 0.9447 [0.9417–0.9476] and 0.8554 [0.8497–0.8611] for the non-imputed models. For MCI→AD, the corresponding values were 0.9020 [0.8947–0.9093] and 0.9185 [0.9128–0.9242] under imputation versus 0.9059 [0.8983–0.9134] and 0.9214 [0.9156–0.9272] without imputation. All differences fell within the estimated confidence intervals, indicating that MICE imputation did not materially alter discriminative performance (Fig S6.1 in S6). Lead-time results were broadly consistent between pipelines (Table 5): detection outcomes, false-alarm rates, and mean lead times were comparable, although for MCI→AD the imputed models detected a smaller fraction of progressors before conversion, reducing the net lead time from 1.64 months to 0.55 months.

Imputation also left the variable rankings largely unchanged (Spearman *ρ* of 0.94 for CN→MCI and 0.88 for MCI→AD between the SHAP shares of the two pipelines). The largest changes involved TOBAC30 and ALCOHOL, whose missingness was associated with the label (S2): under imputation, their MCI→AD shares fell from 5.8% and 2.9% to 0.4% and 0.1%, and TOBAC30, fifth by total gain without imputation, fell to 22nd. Neither variable was recorded at follow-up visits under UDS version 3, and participants enrolled under version 3 progressed less often, so their missing values marked the enrollment period. In the non-imputed MCI→AD models, the 36% of participants with fewer than two recorded TOBAC30 values received a mean SHAP value of −0.28 for TOBAC30, compared with +0.18 for the others. Imputation rounded TOBAC30 and ALCOHOL to their integer codes, which removed this signal without a meaningful change in AUC-ROC.

### Lead-time forecasting

We evaluated the lead-time cohort to characterize how far in advance each classifier could identify future clinical progression and to assess false-alarm rates among non-progressor controls using the optimized decision thresholds and mask lengths established in the methods section. The mean lead time was the average of *t*_conversion_ − *t*_alarm_ over progressors detected before clinical conversion (lead time *>* 0), and the net lead time averaged the same quantity over all detected progressors, including early, at-conversion, and late detections.

Among progressors, the non-imputed CN→MCI models identified progression prior to clinical conversion in 29.58% (95% CI: 28.29%–30.87%) of subjects and at the conversion visit in 60.25% (95% CI: 59.68%–60.82%). Late detection, defined as the first model alarm occurring after conversion, was observed in only 3.28% (95% CI: 2.94%–3.62%) of subjects, while 6.89% (95% CI: 5.88%–7.90%) were not detected. A similar detection profile was observed for the non-imputed MCI→AD models, which identified progression before conversion in 22.41% (95% CI: 21.36%–23.46%) of subjects and at conversion in 53.16% (95% CI: 51.51%–54.82%), with 12.41% (95% CI: 11.29%–13.52%) detected after conversion and 12.03% (95% CI: 10.88%–13.17%) missed. The imputed models produced broadly comparable detection profiles (Fig 6), although for MCI→AD they detected a somewhat smaller fraction of progressors before conversion (20.43% vs. 22.41%) and a larger fraction after conversion (16.91% vs. 12.41%).

**Fig 6.**
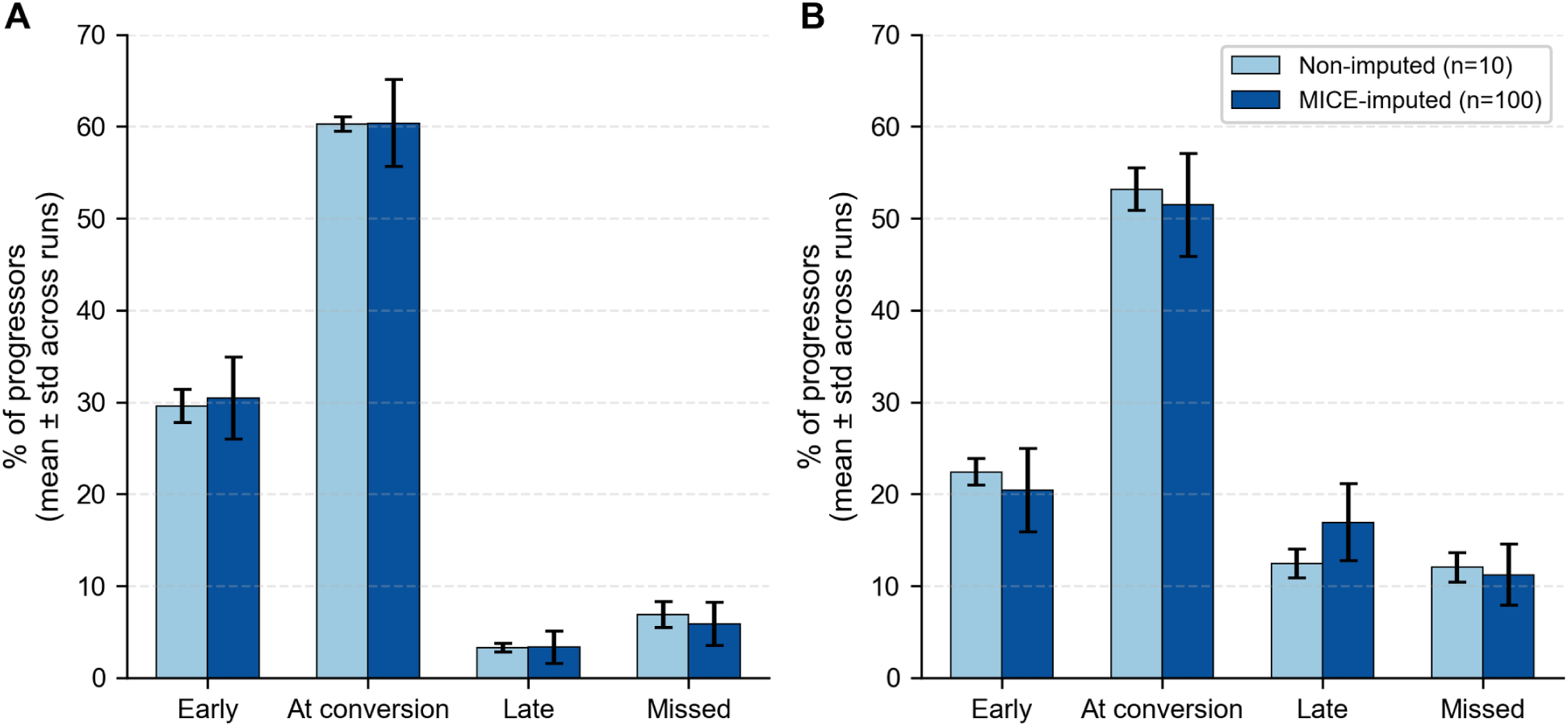
Lead-time detection outcomes for CN MCI and MCI AD forecasting. Percentage of progressors detected early, at the visit of clinical conversion, late (after conversion), or missed entirely. (A) CN→MCI, using a decision threshold of 0.25 and a control mask of three visit intervals. (B) MCI→AD, using a decision threshold of 0.30 and a control mask of two visit intervals. Bars show the mean percentage of progressors in each detection category, averaged across model variants (light = non-imputed models, *n* = 10 runs; dark = MICE-imputed models, *n* = 100 runs); error bars denote ±1 standard deviation across runs.

For the non-imputed models, the mean lead time among subjects detected before clinical conversion was 33.04 months (CN→MCI) and 17.95 months (MCI→AD). Net lead time, averaged across all detected progressors, was 9.92 months (CN→MCI) and 1.64 months (MCI→AD), the latter exceeding the 0.55 months achieved under MICE imputation. Among non-progressor controls, false-alarm rates were 12.6% (CN→MCI) and 20.0% (MCI→AD), compared with 13.2% and 18.1% under MICE imputation, which also yielded similar mean lead times (32.44 and 18.24 months) and a similar CN→MCI net lead time (10.02 months) (Table 5).

## Discussion

We set out to model cognitive trajectories from visit histories of any length, to test whether imputation helps in a cohort with heavy missingness, and to evaluate the resulting models both as classifiers and as early-warning tools. The first two aims had clear answers. Summarizing each variable’s history into a fixed set of temporal descriptors let one gradient-boosted model use every participant’s visits, whether they had two or ten, and it discriminated well in both tasks. Imputation changed discrimination little but changed what the models learned, removing a missingness signal that marked when participants enrolled rather than their clinical state.

The third aim proved the most informative, because classification and early warning are different tasks. A classifier given a participant’s full history has access to the visit at which conversion was recorded; a clinician deciding whether to act sees only the visits made so far. AUC-ROC measures the first and says little about the second. Models that separated progressors from non-progressors well on full histories recognized most progressors only at the visit of diagnosis and warned of a minority beforehand. For a model built on routine records, the size of that minority, how far ahead the warning comes, and how many non-progressors are flagged along the way determine its clinical value.

### Comparison with previous work

Conventional modeling approaches typically rely on cross-sectional baseline snapshots or deep recurrent architectures that require a predetermined number of visits [12, 13, 15, 19, 20]. In contrast, our framework introduces a longitudinal feature-engineering pipeline that collapses variable-length clinical histories into compact temporal descriptors suitable for tree-based learning. Combined with XGBoost’s inherent robustness to missing data and its ability to model complex nonlinear interactions, this design natively accommodates heterogeneous trajectories spanning two to ten visits. This way, we eliminate the need for sequence padding, explicit temporal imputation, or computationally intensive deep sequence models while maintaining high diagnostic performance. Our CN and MCI cohorts were roughly 15 and 6 times larger than those of the two XGBoost studies discussed in the Introduction (12,737 and 2,990 participants, compared with 845 and 493) [12, 16], spanned ages 50 to over 90, and were followed for a mean of 55.6 and 41.3 months, with individual records extending to 231.8 and 203.2 months. Although the cohorts differ and our classifiers use the full visit history rather than a baseline visit, the approach held up in a larger, more heterogeneous cohort followed well beyond prior three- to four-year horizons. This performance strongly suggests that carefully engineered longitudinal features coupled with gradient-boosted decision trees can successfully capture clinically relevant progression patterns while avoiding the greater computational overhead and training resource demands often associated with transformer and recurrent-based architectures.

A core contribution of this work is the introduction of a longitudinal lead-time evaluation framework that shifts the modeling paradigm from static outcome classification to proactive, patient-level tracking. To appreciate how this approach stands out, it is helpful to contrast it with existing clinical paradigms. As synthesized in a comprehensive review in 2025, contemporary dynamic prediction of Alzheimer’s disease is dominated by four main mathematical frameworks: two-stage models, joint models, landmark models, and deep learning architectures [46]. While these conventional paradigms excel at updating population-level survival probabilities or predicting a discrete time-to-event as new clinical information arrives, they are generally evaluated in terms of updated survival probabilities or event risks rather than the timing and stability of discrete warning alerts generated for individual patients. Similarly, even advanced machine learning benchmarks treat time as a predefined prediction horizon rather than an outcome to be estimated. For example, a recent preprint by Farzana et al. evaluated Alzheimer’s disease and related dementia onset prediction at four fixed lead times of 6, 12, 24, and 36 months prior to diagnosis using longitudinal electronic health records [47]. While this design provides useful insight into discriminative performance at clinically meaningful cutoffs, it does not track the emergence of a predictive signal over time for individual patients, nor does it characterize the distribution of lead times actually achieved across the population.

Our framework reframes the evaluation problem away from both fixed-horizon prediction and conventional time-to-event modeling toward individualized threshold-based tracking. By sequentially truncating clinical histories visit-by-visit, we emulate a deployment scenario in which risk estimates are updated whenever new clinical information becomes available. Rather than evaluating performance at a single predefined horizon, we track subject-specific risk trajectories and quantify the timing of the first threshold crossing. This approach directly yields mean lead time, net lead time, and longitudinal false-alarm burden—quantities that fixed-horizon evaluations cannot provide. To our knowledge, this is among the first frameworks to evaluate non-imaging Alzheimer’s progression models through explicit measurement of patient-level lead times and longitudinal false-alarm burden.

### Feature attribution

The two tasks relied on different information. The CN→MCI model depended most on CDR-SB and gave more weight than the MCI→AD model to non-cognitive markers: a lower BMI and rising depressive symptoms raised predicted risk, in line with the lower BMI observed in the decade before a dementia diagnosis [48] and with depression as a risk factor for dementia [29]. The MCI→AD model gave more weight to the FAQ items and the Community Affairs score, consistent with the loss of independence in daily activities that distinguishes dementia from MCI. Pang et al. likewise found CDR subitems, including community affairs, among the main predictors of conversion to AD within three years in NACC [19]. In contrast with CN→MCI, higher GDS scores lowered the predicted risk of AD. Because the GDS is self-reported, this may reflect reduced awareness of symptoms, which predicts conversion from MCI to AD [49], or MCI caused by depression rather than neurodegeneration. Two caveats apply to these attributions. CDR-SB and the FAQ are part of the information clinicians use to assign the diagnoses that define our labels, so some of their weight reflects this overlap rather than independent prognostic value. The engineered features are also correlated, so how attribution is divided among them is partly arbitrary. With these caveats, trajectory features received about as much attribution as level features for CN→MCI and more for MCI→AD, and the change in CDR-SB since the first visit ranked among the four most important features in both tasks, suggesting that the longitudinal summaries added information beyond the most recent visit. Lagged values, acceleration, and the hearing–vision interaction terms added little.

The comparison of pipelines shows that the non-imputed models also used missingness as a predictor. XGBoost learns a default branch for missing values at each split, and exploiting missingness in this way can improve accuracy [25]. In NACC, however, some missingness reflects changes to the UDS rather than the participant’s clinical state: participants enrolled under version 3 had no follow-up tobacco or alcohol items, had shorter follow-up, and were less often observed to progress (S2). Version 3 also replaced the MMSE with the MoCA; converting MoCA scores to the MMSE scale [35] left only 2% of participants without a cognitive screening score and largely removed that source of enrollment-dependent missingness. The tobacco and alcohol items still marked the enrollment period in the non-imputed models, and imputation removed this signal without a meaningful change in AUC-ROC. Discrimination was nonetheless lower for participants enrolled under version 3, who were followed for a shorter time: AUC-ROC was 0.909 versus 0.960 for earlier enrollees in the CN→MCI task and 0.801 versus 0.909 in the MCI→AD task (S2). Restricting analyses to a single UDS version, as Pang et al. did [19], would avoid this heterogeneity.

### Ethics

The ethical implications of a large-scale statistical learning-based approach to predicting dementia must be considered. Firstly, a system could introduce a false level of certainty in diagnosis, resulting in a false negative case where dementia is undetected. This uncertainty exists with physician diagnoses also, but growing confidence in systems relying on artificial intelligence may obscure this risk. Systems which use predictive modeling to assess risk for developing dementia may have unintended side effects. Healthcare providers could deny older adults coverage based on a set of behavioral or genetic risk factors. Employers could deny work to older adults who increasingly find themselves unable to retire. Work must be done to ensure these diagnostic instruments for detection of AD are used in an equitable and responsible way.

### Limitations and future research

The NACC dataset is a convenience sample of patients who present to Alzheimer’s Disease Research Centers and may not be representative of the general population. We chose to use non-imaging clinical data to make the model more accessible and cost-effective, but this choice may limit the model’s performance relative to imaging-based approaches. MRI studies relating to AD detection should be incorporated as an improved diagnostic measurement with an acceptable level of accessibility to the general population. Future work should explore how to integrate imaging data into our longitudinal framework to improve performance.

The use of MCI as a ground truth label is also a limitation, as MCI is a broadly defined condition with multiple etiologies. Not all individuals with MCI will progress to AD. This heterogeneity may introduce noise into the model’s predictions and affect its generalizability. Future research should consider more specific biomarkers or clinical criteria to define progression to AD.

Converting MoCA scores to the MMSE scale also adds measurement error. The crosswalk maps every MoCA score from 27 to 30 to an MMSE of 30 [35], so converted scores lose sensitivity near the ceiling, where many cognitively normal participants score.

Finally, because a participant is labeled a progressor only when a more advanced visit is observed, progressors in the MCI→AD cohort had more visits and longer follow-up than controls (median 4 vs. 3 visits; 42.1 vs. 24.2 months), which could let a model learn follow-up length rather than clinical change. Removing the features that directly encode visit timing and visit count changed AUC-ROC by less than 0.01. When every participant was restricted to their first two visits, AUC-ROC was 0.842. After excluding progressors who had converted by their second visit, it was 0.797, compared with 0.929 for the full-history model on the same participants. The full-history model’s discrimination of later converters therefore relies partly on visits after the second, which include the conversion visit (see S5).

## Conclusion

Using only routinely collected, non-imaging clinical data, gradient-boosted trees trained on variable-length visit histories distinguished progressors from non-progressors with AUC-ROCs above 0.90 in both tasks.

Applied visit by visit, the models flagged 29.6% of CN→MCI progressors a mean of 33 months before diagnosis and 22.4% of MCI→AD progressors a mean of 18 months before, with false-alarm rates of 12.6% and 20.0%. Such models could help decide who is referred for costlier biomarker testing. Validation in external and prospective cohorts is the next step toward using such models for low-cost risk stratification.

## Code availability

All code used for preprocessing, feature engineering, model training, and lead-time analysis is publicly available at https://github.com/Andrew-Gautier/XGBoost-AD-Forecasting.

## Data Availability

No data were generated by this study. The analyses used existing, restricted-access data from the National Alzheimer's Coordinating Center (NACC) Uniform Data Set and Neuropathology (NP) Form, December 2025 Data Freeze. The authors received these data under a NACC Data Use Agreement and are not permitted to redistribute individual-level records; NACC serves as the third-party repository and provides the identical dataset to any qualified investigator who submits a data request. The authors had no special access privileges that other researchers would not have.

https://www.naccdata.org/data-request-process

## Acknowledgments

Research reported in this paper was supported by the National Institute on Aging of the National Institutes of Health under Award Number R25AG059558 through the AGE-ADAR Scholars Program at West Virginia University.

The NACC database is funded by NIA/NIH Grant U24 AG072122. NACC data are contributed by the NIA-funded ADRCs: P30 AG062429 (PI James Brewer, MD, PhD), P30 AG066468 (PI Oscar Lopez, MD), P30 AG062421 (PI Teresa Gomez-Isla, MD), P30 AG066509 (PI Thomas Grabowski, MD), P30 AG066514 (PI Mary Sano, PhD), P30 AG066530 (PI Helena Chui, MD, Arthur Toga, PhD), P30 AG066507 (PI Marilyn Albert, PhD), P30 AG066444 (PI David Holtzman, MD), P30 AG066518 (PIs Lisa Silbert, MD, Kevin Duff, PhD), P30 AG066512 (PI Thomas Wisniewski, MD), P30 AG066462 (PI Scott Small, MD), P30 AG072979 (PI David Wolk, MD), P30 AG072972 (PIs Charles DeCarli, MD, Rachel Whitmer, PhD), P30 AG072976 (PI Andrew Saykin, PsyD), P30 AG072975 (PI Julie Schneider, MD, MS), P30 AG072978 (PI Ann McKee, MD), P30 AG072977 (PI Robert Vassar, PhD), P30 AG066519 (PI Joshua Grill, PhD), P30 AG062677 (PIs Brad Boeve, MD, Ronald Petersen, MD, PhD), P30 AG079280 (PI Jessica Langbaum, PhD), P30 AG062422 (PI Gil Rabinovici, MD), P30 AG066511 (PI Allan Levey, MD, PhD), P30 AG072946 (PI Linda Van Eldik, PhD), P30 AG062715 (PI Sanjay Asthana, MD, FRCP), P30 AG072973 (PI Russell Swerdlow, MD), P30 AG066506 (PIs Glenn Smith, PhD, ABPP, David Lowenstein, PhD, Ranjan Duara, MD), P30 AG066508 (PIs Stephen Strittmatter, MD, PhD, Christopher Van Dyck, MD), P30 AG066515 (PI Victor Henderson, MD, MS), P30 AG072947 (PI Suzanne Craft, PhD), P30 AG072931 (PI Henry Paulson, MD, PhD), P30 AG066546 (PIs Sudha Seshadri, MD, Gladys Maestre, MD, PhD), P30 AG086401 (PI Erik Roberson, MD, PhD), P30 AG086404 (PI Gary Rosenberg, MD), P30 AG086403 (PI Angela Jefferson, PhD), P30 AG072958 (PIs Heather Whitson, MD, Gwenn Garden, MD, PhD), P30 AG072959 (PI Jagan Pillai, MD, PhD), P30 AG092752 (Ihab Hajjar, MD, MS).

## Supplementary material

### S1 Variables used from the NACC dataset

Definitions and coding of all static and longitudinal variables, including the hearing and vision composite encoding.

Table S1.1 lists the 17 static variables and Table S1.2 the 20 longitudinal variables and the time reference months since baseline. Descriptions follow the NACC Uniform Data Set definitions. Each static variable takes the participant’s most recent recorded value; visits at which an item was not collected, such as the health history at UDS version 3 follow-up visits, are skipped. Binary comorbidities were recoded so that 1 indicates an active or recent condition and 0 an absent, remote/inactive, or unknown one. The ten FAQ items record whether, in the past four weeks, the participant had difficulty or needed help with the stated activity (0 = normal to 3 = dependent).

**Table S1.1.** Static variables.

| Variable | Type | Description |
| --- | --- | --- |
| age | Continuous | Age at initial visit (NACCAGEB), years |
| EDUC | Continuous | Years of education |
| SMOKYRS | Continuous | Total years smoked cigarettes (0–87; 88 = not applicable and 99 = unknown set to missing) |
| NACCFAM | Binary | First-degree family member with cognitive impairment |
| CVHATT | Binary | Heart attack/cardiac arrest |
| CVAFIB | Binary | Atrial fibrillation |
| DIABETES | Binary | Diabetes |
| HYPERCHO | Binary | Hypercholesterolemia |
| HYPERTEN | Binary | Hypertension |
| B12DEF | Binary | Vitamin B<sub>12</sub> deficiency |
| DEPD | Binary | Depression or dysphoria in the last month |
| ANX | Binary | Anxiety in the last month |
| NACCTBI | Binary | History of traumatic brain injury |
| HISPANIC | Binary | Hispanic/Latino origin |
| SEX | Categorical | Sex |
| RACE | Categorical | Race |
| NACCNE4S | Categorical | Number of APOE ε4 alleles (0/1/2) |

**Table S1.2.** Longitudinal variables. Ranges are those retained after sentinel codes were set to missing.

| Variable | Range | Description |
| --- | --- | --- |
| *Cognitive and psychiatric scales* |  |  |
| NACCM MSE | 0–30 | Total MMSE score; at UDS version 3 visits without an MMSE, the education-adjusted MoCA score (NACCMOCA) converted to the MMSE scale (see Methods); lower = worse |
| CDRSUM | 0–18 | CDR<sup>®</sup> Sum of Boxes; higher = worse |
| NACCGDS | 0–15 | Total GDS score |
| *Functional Activities Questionnaire (FAQ) items* |  |  |
| BILLS | 0–3 | Writing checks, paying bills, or balancing a checkbook |
| TAXES | 0–3 | Assembling tax records, business affairs, or other papers |
| SHOPPING | 0–3 | Shopping alone for clothes, household necessities, or groceries |
| GAMES | 0–3 | Playing a game of skill such as bridge or chess, or working on a hobby |
| STOVE | 0–3 | Heating water, making a cup of coffee, or turning off the stove |
| MEALPREP | 0–3 | Preparing a balanced meal |
| EVENTS | 0–3 | Keeping track of current events |
| PAYATTN | 0–3 | Paying attention to and understanding a TV program, book, or magazine |
| REMDATES | 0–3 | Remembering appointments, family occasions, holidays, or medications |
| TRAVEL | 0–3 | Traveling out of the neighborhood, driving, or arranging public transportation |
| *Physical health and behavior* |  |  |
| NACCBMI | 10–100 | Body mass index |
| TOBAC30 | 0/1 | Smoked cigarettes in the last 30 days |
| ALCOHOL | 0/1/2 | Alcohol abuse: clinically significant impairment over a 12-month period in work, driving, legal, or social domains (0 = absent, 1 = recent/active, 2 = remote/inactive; recoded before feature engineering, see S3) |
| *Social and sensory* |  |  |
| NACCLIVS | 1–5 | Living situation, nominal: 1 = alone, 2 = with spouse or partner, 3 = with relative or friend, 4 = with group, 5 = other |
| COMMUN | 0–3 | CDR Community Affairs domain score (0, 0.5, 1, 2, 3) |
| hearing | 0–2 | Composite of HEARING, HEAR AID, and HEARWAID (Fig S1.1) |
| vision | 0–2 | Composite of VISION, VISCORR, and VISWCORR (Fig S1.1) |
| *Time reference* |  |  |
| months_since_baseline | — | Months since the initial visit, to two decimal places; derived from NACCFDYS |

**Fig S1.1.**
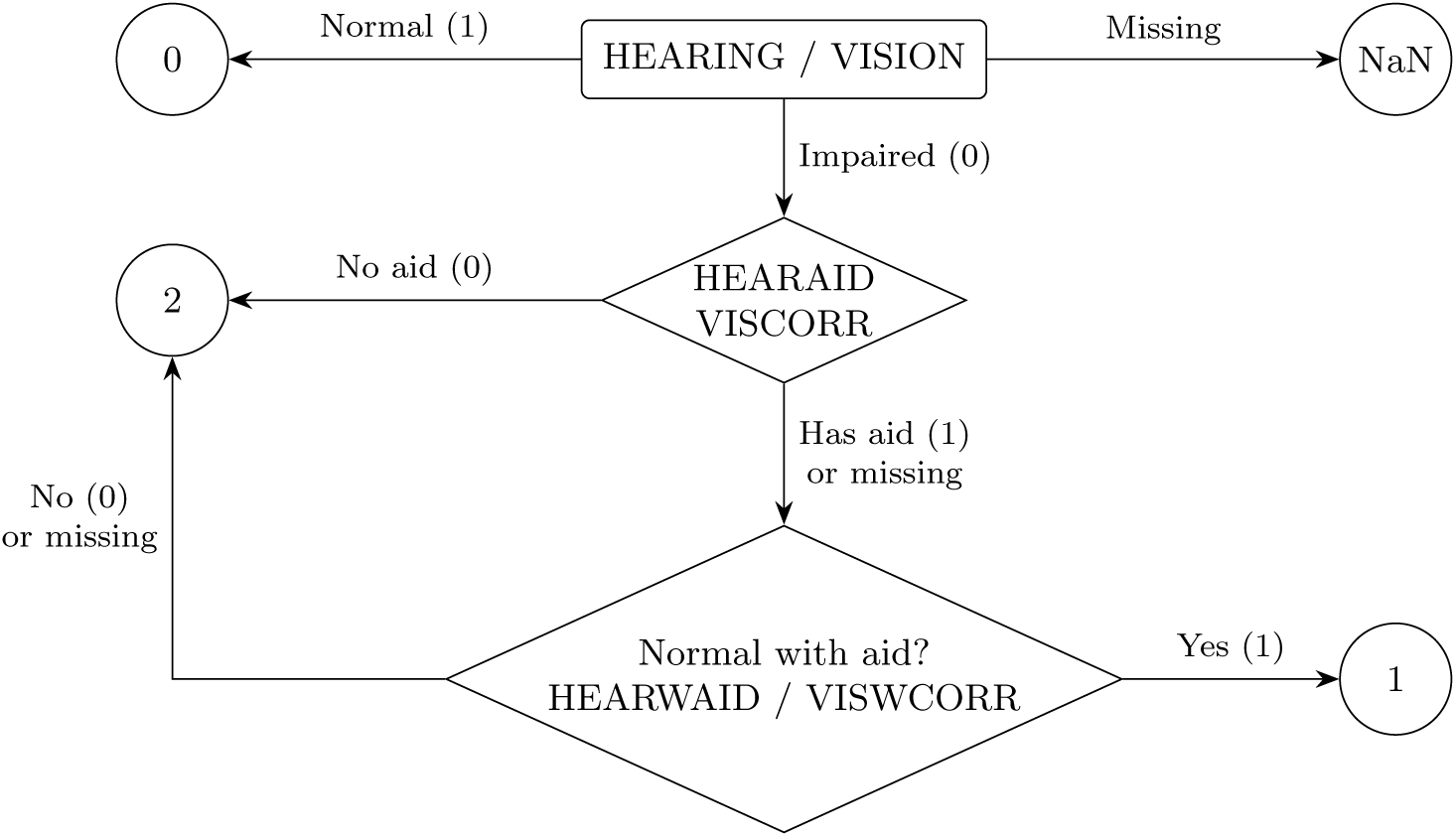
Encoding of the hearing and vision composites. At each visit, the base indicator (HEARING or VISION) is read first: functionally normal is coded 0 and missing remains missing. If impairment is present, the presence of an assistive device (HEARAID or VISCORR) and whether function is normal with it (HEARWAID or VISWCORR) determine the code: 1 = impaired but corrected with a device, 2 = impaired without effective correction.

#### S2 Missingness analysis

Patterns of missing data by cohort and progression status, co-missingness across variables, and missingness across visits.

### Methods

Missingness was assessed separately for the CN→MCI and MCI→AD cohorts, after sentinel codes were set to missing. For each longitudinal variable, we computed the proportion of each participant’s visits at which the variable was missing, and report the mean of this proportion across participants with a 95% percentile bootstrap confidence interval (2,000 resamples of participants), by progression status (Fig S2.1). For static variables, measured once per participant, missingness is a binary indicator. We also computed the correlation of the per-participant missingness proportions between longitudinal variables (Fig S2.2) and the proportion of participants missing each variable at each visit number (Fig S2.3).

### Results

Only two static variables had missing values: NACCNE4S, missing for 14–24% of participants and more often in non-progressors, and SMOKYRS, missing for fewer than 3%. All other static variables were complete.

Among longitudinal variables, NACCMOCA was the most often missing, because the MoCA was administered only under UDS version 3, followed by TOBAC30 and ALCOHOL (Fig S2.1). After MoCA scores were converted to the MMSE scale, NACCMMSE was missing at 10–14% of a participant’s visits in both cohorts and progression groups, compared with 27–63% before conversion. CDRSUM and COMMUN were complete and NACCLIVS nearly so. For TOBAC30 and ALCOHOL, non-progressors had more missing visits than progressors, most markedly in the MCI→AD cohort (35% vs. 16%), whereas NACCMOCA was missing more often in progressors, who more often enrolled before version 3.

Missingness co-occurred within domains (Fig S2.2). The FAQ items formed a correlated cluster, hearing and vision were almost always missing together (*r* ≥ 0.93), and TOBAC30 and ALCOHOL were missing together (*r* ≥ 0.99). NACCMOCA missingness was inversely correlated with that of TOBAC30 (*r* = −0.77 and −0.87), because both depend on the UDS version. Missingness of TOBAC30, ALCOHOL, NACCBMI, and NACCMMSE rose across successive visits, whereas NACCMOCA missingness was lowest at the latest visits (Fig S2.3). TOBAC30 and ALCOHOL were missing at fewer than 1% of first visits but for over 90% of participants at the tenth.

### Missingness and UDS version

NACC revised the Uniform Data Set (UDS) during the study period. Of the 14,925 participants used to train and test the classifiers, 4,473 enrolled under version 1, 3,687 under version 2, and 6,765 under version 3. Version 3 replaced the MMSE with the Montreal Cognitive Assessment (MoCA) [1]: the MMSE was missing at more than 95% of version 3 visits, compared with 4–11% of earlier visits, while the MoCA was recorded at 94–95% of version 3 initial visits and 72–77% of follow-up visits. Converting MoCA scores to the MMSE scale (see Methods) left 2.0–2.5% of participants without a cognitive screening score at any visit, compared with 34–50% before conversion. TOBAC30 and ALCOHOL were missing at every version 3 follow-up visit and at fewer than 1% of other visits. Participants enrolled under version 3 had shorter follow-up (median 37 vs. 52–62 months in the CN→MCI cohort and 25 vs. 34–37 months in the MCI→AD cohort) and progressed less often: 11.5%, 20.8%, and 27.2% of CN participants enrolled under versions 3, 2, and 1 progressed, as did 24.7%, 45.5%, and 69.4% of MCI participants. The UDS version at enrollment alone discriminated progressors with an AUC-ROC of 0.62 (CN→MCI) and 0.71 (MCI→AD). The label-associated missingness of TOBAC30 and ALCOHOL therefore tracks the enrollment period, as did that of the MMSE before MoCA scores were converted. Discrimination also differed by enrollment period: in the test sets of the non-imputed models (means of ten runs), AUC-ROC was 0.960 for participants enrolled under versions 1–2 and 0.909 for those enrolled under version 3 in the CN→MCI task, and 0.909 and 0.801 in the MCI→AD task.

**Fig S2.1.**
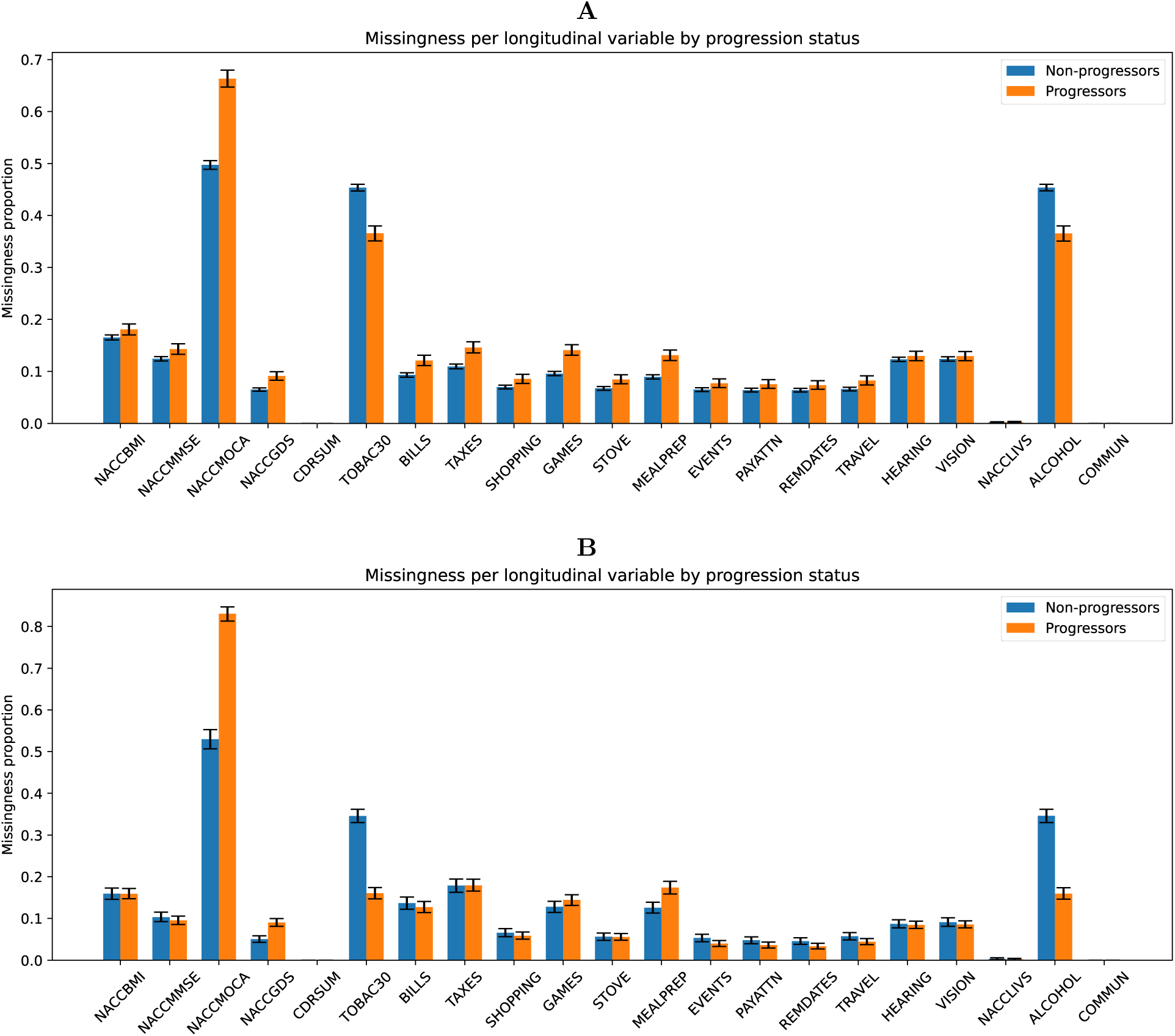
Missingness of longitudinal variables by progression status. (A) CN→MCI and (B) MCI→AD cohorts. Bars show the mean per-participant proportion of missing visits; error bars show 95% percentile bootstrap confidence intervals of the mean.

**Fig S2.2.**
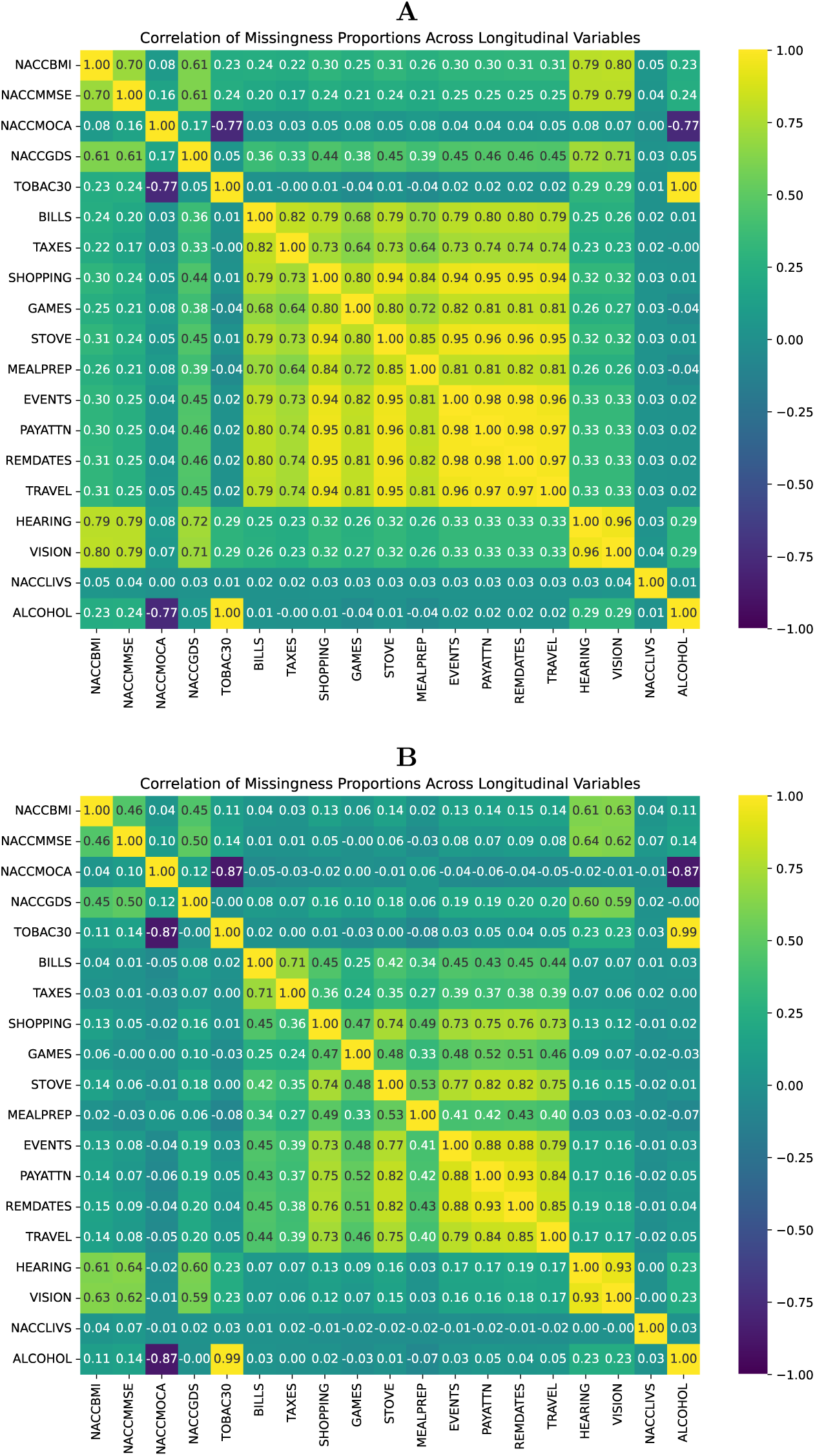
Correlation of missingness between longitudinal variables. (A) CN→MCI and (B) MCI→AD cohorts. Each cell is the correlation, across participants, of the per-participant proportions of missing visits for two variables. CDRSUM and COMMUN are omitted because they are never missing.

**Fig S2.3.**
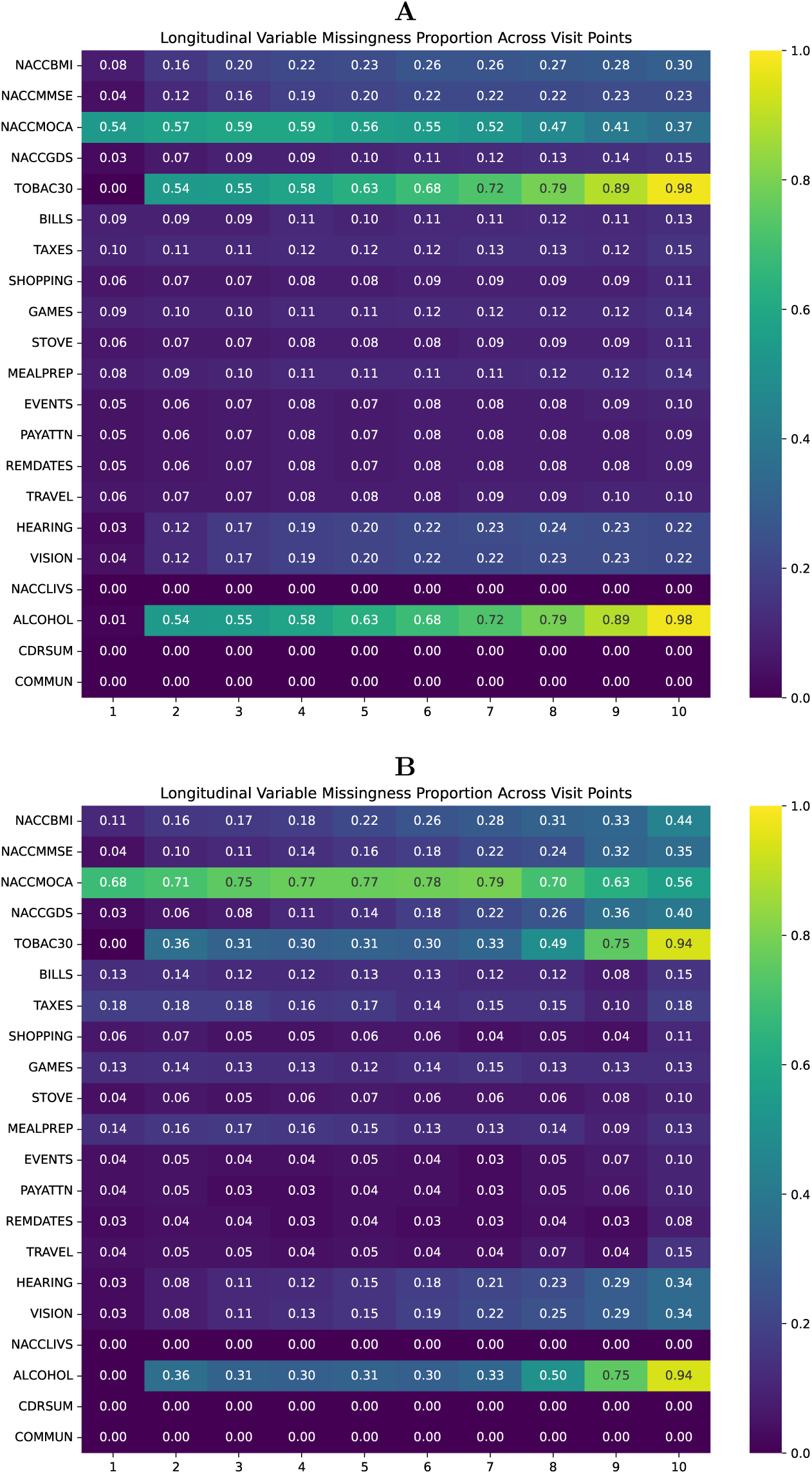
Missingness of longitudinal variables across visits. (A) CN→MCI and (B) MCI→AD cohorts. Each cell is the proportion of participants with that visit whose value is missing.

## S3 Feature engineering

Full description of the engineered feature set.

### Overview

The feature engineering pipeline transforms per-subject longitudinal visit records into a fixed-width, visit-count-agnostic feature vector suitable for gradient-boosted tree classification. Each subject has between 2 and 10 visits, and repeated measurements are stored as variable-length lists (one entry per visit) that cannot be passed directly to a tabular classifier. The pipeline extracts summary statistics from these lists, yielding a single row per subject regardless of the number of visits observed. The transformation is implemented in create delta features() and preprocess data(), and is applied independently to the training and test partitions of each split, so no information leaks from held-out samples.

The final feature set contains four categories:

1. **Static features**: demographics and comorbidities measured once or assumed stable across visits (17 variables).
2. **Longitudinal summary features**: visit-agnostic statistics derived from each numeric repeated-measurement variable (up to 13 statistics per variable).
3. **Living-situation features**: the first and last recorded codes of the nominal NACCLIVS and a flag for any change (3 features).
4. **Interaction features**: explicit terms capturing dual sensory impairment (3 features).

### Input variables

The 17 static and 20 longitudinal input variables are defined in Table S1.1 and Table S1.2. Static variables are passed to the model without transformation. NACCMMSE holds the MMSE score or, at visits without one, the education-adjusted MoCA score converted to the MMSE scale with the NACC crosswalk [1]. Each longitudinal variable is stored as a list of per-visit measurements, with missing visits represented as NaN. The hearing and vision composites range from 0 (no impairment) to 2 (impaired without effective aid) (Fig S1.1). ALCOHOL is recoded before feature extraction (0 = absent → 0, 2 = remote/inactive → 1, 1 = recent/active → 2) so that higher values always indicate greater clinical concern.

### Longitudinal feature engineering

Let V(*x*) = {*x_i_* : *x_i_* ≠ NaN} denote the valid (non-missing) observations of variable *x*, ordered chronologically, and let *n* = |V(*x*)| be the number of valid visits. Where time is referenced, *t_i_* denotes the months since baseline of the *i*-th valid observation.

#### Summary statistics

Nine statistics are computed for every numeric longitudinal variable, that is, all except NACCLIVS:

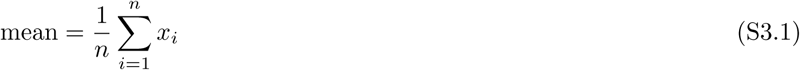

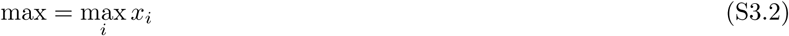

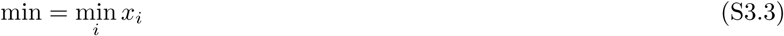

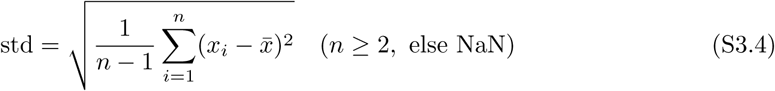

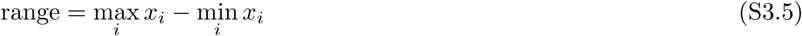

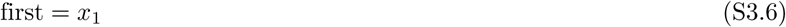

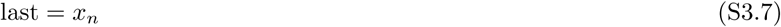

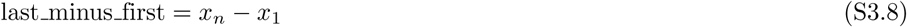

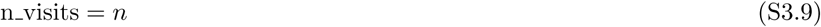

#### Time-normalized slope

Slopes are computed against elapsed time in months rather than visit index, so they are expressed in clinical units per month and are comparable across patients with irregular visit spacing. The slope is estimated by ordinary least squares (OLS):

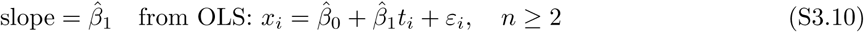

When fewer than 2 valid observations exist, slope is set to NaN.

#### Time-normalized acceleration

Acceleration captures the rate of change in the rate of change, i.e., whether a patient is accelerating into decline. Define the pairwise per-month slopes between consecutive valid visits:

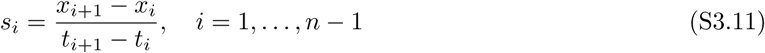

Acceleration is the OLS slope of *s_i_* regressed on the midpoint times *τ_i_* = (*t_i_* + *t_i_*_+1_)*/*2:

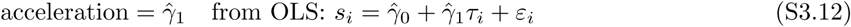

Acceleration is set to NaN for any patient with fewer than 4 valid observations, so that the regression uses at least 3 pairwise slopes. This threshold was raised from 3 to 4 valid visits to mitigate the instability of second-derivative estimates from very short sequences.

#### Slope variability

A single linear slope cannot capture non-linear trajectories, step-changes, or periods of temporary recovery. The standard deviation of the pairwise per-month slopes quantifies this variability:

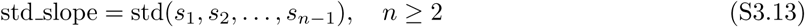

where *s_i_*is defined in Eq (S3.11) and std(·) is the sample standard deviation (ddof = 1). When *n* = 2, exactly one slope exists and std slope = 0; when *n <* 2, std slope is NaN.

#### Maximum percentage change

Alzheimer’s progression can involve discrete step-changes following clinical events. The maximum relative change between consecutive visits captures this:

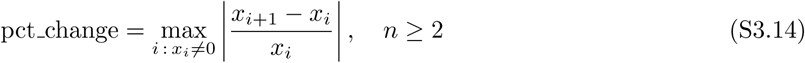

Pairs with *x_i_* = 0 are excluded to prevent division by zero. When every consecutive pair has a zero denominator, or *n <* 2, the feature is NaN.

#### Lagged observations

Summary statistics such as mean and slope discard temporal ordering: two patients with the same mean and slope but different trajectories (improved then worsened vs. worsened then improved) are indistinguishable. Lagged features resolve this by encoding the most recent values explicitly:

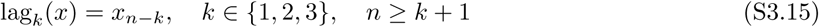

Thus lag1, lag2, and lag3 are the second-, third-, and fourth-to-last valid observations, and are NaN when too few valid observations exist. Lags are computed for seven clinically prioritized variables (NACCMMSE, CDRSUM, NACCGDS, hearing, vision, BILLS, and SHOPPING), yielding 21 features. These cover the primary cognitive screening instrument, global disease severity, depression, both sensory modalities, and two representative functional items.

#### Living situation

NACCLIVS is nominal (1 = alone, 2 = with spouse or partner, 3 = with relative or friend, 4 = with group, 5 = other), so arithmetic summaries of its codes are meaningless: a mean of 1 and 3, for example, is not “with spouse or partner.” It contributes three features instead: its first and last recorded codes, treated as categorical, and NACCLIVS changed, which is 1 if the recorded code differs between any two visits and 0 otherwise (NaN if never recorded). NACCLIVS is not imputed.

#### Visit timing features

Because months since baseline is the time axis itself, a slope or acceleration over it is not meaningful. Instead, two features describe the spacing of visits:

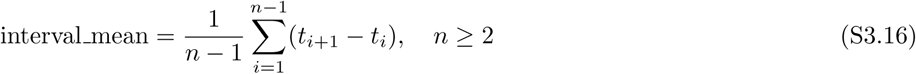

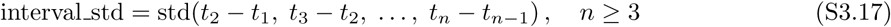

interval mean is the average time between assessments (months), and interval std captures the irregularity of the visit schedule. The nine summary statistics are also computed for months since baseline, so that features such as months since baseline last (total follow-up duration) are available to the model.

#### Hearing × vision interaction features

Dual sensory impairment, concurrent hearing and vision loss, is a well-documented risk factor for dementia, and its effect is reported to be multiplicative rather than additive [2]. A model that treats hearing and vision as independent inputs cannot capture this synergy, so three interaction terms are computed from the last valid value of each composite:

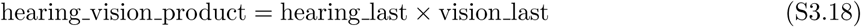

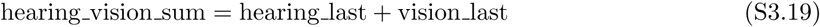

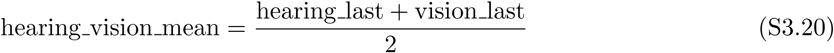

The product is zero if either sense is unimpaired and positive only when both are impaired, directly encoding co-occurrence. The sum and mean capture overall sensory burden.

#### Feature summary

Table S3.1 gives the number of features in each category, and Table S3.2 lists the features created for each numeric longitudinal variable together with the minimum number of valid visits each requires.

**Table S3.1.** Number of features by category. Final feature matrix passed to the XGBoost classifier.

| Category | Per variable | Total |
| --- | --- | --- |
| Static (numeric, binary, categorical) | 1 | 17 |
| Longitudinal summary (19 variables × 11 statistics) | 11 | 209 |
| Standard summary | 9 | 171 |
| Time-normalized slope | 1 | 19 |
| Acceleration ($\geq 4$ visits) | 1 | 19 |
| Slope variability, std_slope (19 variables) | 1 | 19 |
| Maximum percentage change, pct_change (19 variables) | 1 | 19 |
| Living situation (first, last, changed) | — | 3 |
| Visit timing (months_since_baseline) | — | 11 |
| Standard summary | — | 9 |
| Interval mean and std | — | 2 |
| Lagged (7 variables × 3 lags) | 3 | 21 |
| Hearing × vision interactions | — | 3 |
| **Total** |  | **302** |

#### Design rationale

##### Visit-count independence

Because patients have differing numbers of visits, all features are summary statistics that collapse the visit dimension. The model therefore generalizes across patients with 2 and 10 visits without imputation or fixed-length padding.

##### Time normalization

All rate features (slope, acceleration, std slope) use elapsed months rather than visit index. A 10-point MMSE decline over 6 months is therefore distinguished from the same decline over 4 years, as the former indicates a much more aggressive trajectory.

##### Missing values

Features that require more observations than a patient has (e.g., acceleration requires ≥ 4) are set to NaN rather than imputed. XGBoost handles missing values natively through a learned default branch direction at each split.

##### No scaling

No standardization or normalization is applied. XGBoost is scale-invariant, and omitting scaling removes any possibility of test-set information influencing training-set normalization.

**Table S3.2.**
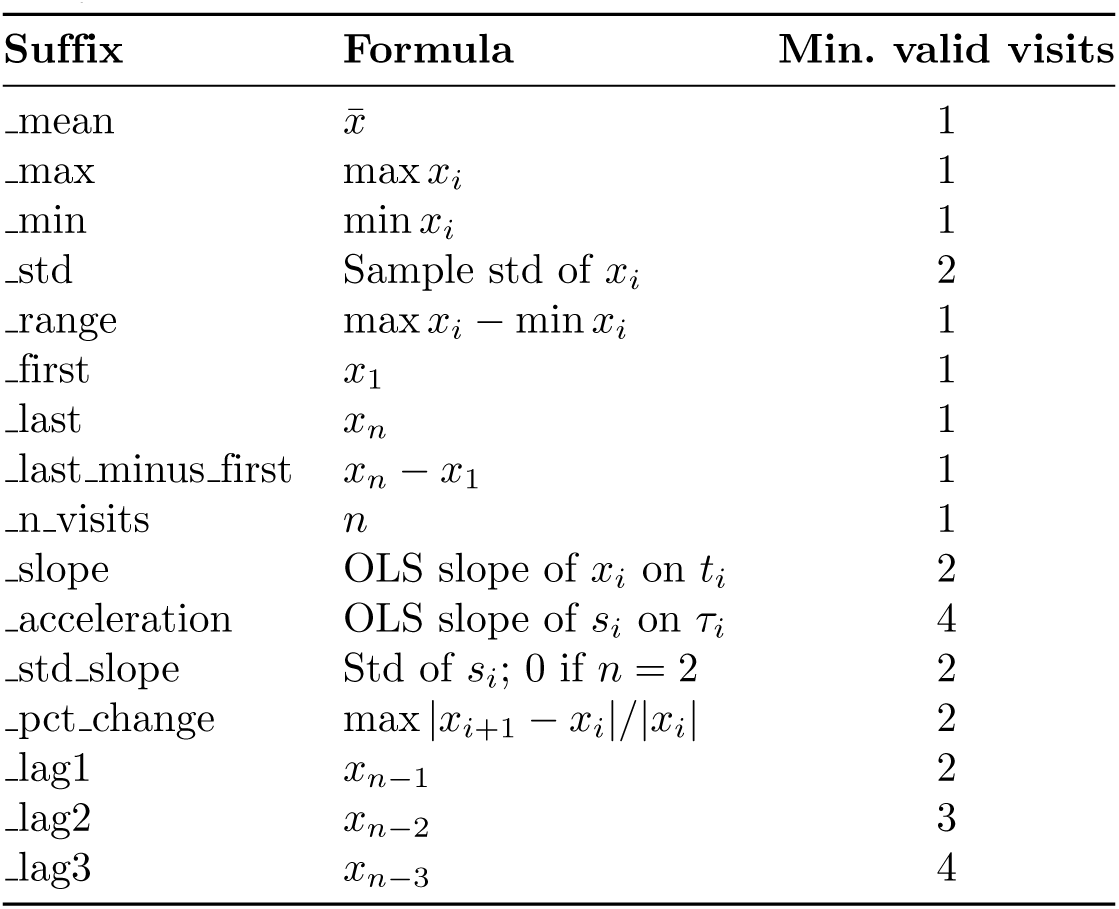
Features created for each numeric longitudinal variable. Lags are computed for the seven selected variables only.

##### Categorical encoding

The five categorical features (SEX, RACE, NACCNE4S, NACCLIVS first, and NACCLIVS last) are declared categorical to XGBoost through its feature types, so trees split them by groups of categories rather than by numeric code, which is appropriate for unordered factors. NACCNE4S (the APOE *ε*4 allele count) is ordinal, but with only three levels, any split on its numeric order is also a split by groups of categories, so treating it as categorical loses nothing.

##### Alcohol severity recode

NACC codes alcohol use as 0 = absent, 1 = recent/active, and 2 = remote/inactive, an ordering in which recent use is numerically lower than remote use. ALCOHOL is therefore recoded to 0/2/1 before summary and slope features are computed, so that larger values and positive slopes consistently indicate greater use.

##### No data leakage

create delta features() and preprocess data() are called independently on the training and test partitions of each split. No statistics (means, scalers, imputers) are fit on combined training and test data.

##### Target variable

The binary label Prog ID is derived from each subject’s sequence of visit-level diagnoses *d*_1_*, . . ., d_m_*, coded 0 = CN, 1 = MCI, and 2 = AD:

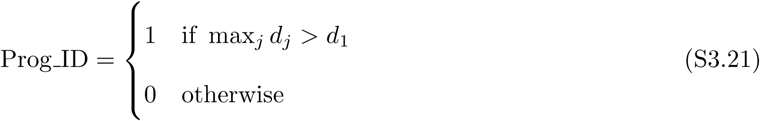

For the CN→MCI cohort, Prog ID = 1 if the subject ever progressed beyond cognitively normal. For the MCI→AD cohort, Prog ID = 1 if the subject was ever diagnosed with AD.

#### S4 Lead-time experiment

Lead-time cohort composition, sequence truncation and masking, and selection of decision thresholds and mask lengths.

#### Lead-time cohort

The lead-time holdout was built by sampling 5% of participants from each of four groups (CN→MCI progressors and non-progressors, MCI→AD progressors and non-progressors), preserving the class structure of the full dataset. Non-imputed and MICE-imputed versions contain the same participants and follow identical downstream processing. Table S4.1 summarizes the cohort.

**Table S4.1.** Demographics of the lead-time cohort. Mean ± SD or *n* (%).

| | All<br>($N = 802$) | Stable CN<br>($n = 526$) | CN→MCI<br>($n = 119$) | Stable MCI<br>($n = 78$) | MCI→AD<br>($n = 79$) |
| --- | --- | --- | --- | --- | --- |
| Observation period (months) | 53.45 ± 37.11 | 53.55 ± 38.32 | 67.17 ± 36.52 | 35.26 ± 26.08 | 50.04 ± 30.68 |
| Visit interval (months) | 15.94 ± 6.25 | 15.87 ± 5.76 | 16.98 ± 9.26 | 16.01 ± 5.00 | 14.83 ± 4.44 |
| Age (years) | 71.34 ± 9.02 | 69.92 ± 8.57 | 75.90 ± 8.49 | 68.56 ± 8.33 | 76.66 ± 9.01 |
| *Age group* |  |  |  |  |  |
| 50–60 | 83 (10.3) | 65 (12.4) | 3 (2.5) | 12 (15.4) | 3 (3.8) |
| 61–70 | 317 (39.5) | 232 (44.1) | 30 (25.2) | 37 (47.4) | 18 (22.8) |
| 71–80 | 262 (32.7) | 160 (30.4) | 49 (41.2) | 23 (29.5) | 30 (38.0) |
| 81–90 | 127 (15.8) | 64 (12.2) | 34 (28.6) | 5 (6.4) | 24 (30.4) |
| >90 | 13 (1.6) | 5 (1.0) | 3 (2.5) | 1 (1.3) | 4 (5.1) |
| *Sex* |  |  |  |  |  |
| Male | 305 (38.0) | 196 (37.3) | 37 (31.1) | 37 (47.4) | 35 (44.3) |
| Female | 497 (62.0) | 330 (62.7) | 82 (68.9) | 41 (52.6) | 44 (55.7) |
| *Race* |  |  |  |  |  |
| White | 612 (76.3) | 397 (75.5) | 95 (79.8) | 59 (75.6) | 61 (77.2) |
| Black or African American | 148 (18.5) | 99 (18.8) | 19 (16.0) | 15 (19.2) | 15 (19.0) |
| American Indian or Alaska Native | 8 (1.0) | 4 (0.8) | 2 (1.7) | 1 (1.3) | 1 (1.3) |
| Native Hawaiian or Other Pacific Islander | 0 (0.0) | 0 (0.0) | 0 (0.0) | 0 (0.0) | 0 (0.0) |
| Asian | 22 (2.7) | 18 (3.4) | 1 (0.8) | 2 (2.6) | 1 (1.3) |
| Other or unknown | 12 (1.5) | 8 (1.5) | 2 (1.7) | 1 (1.3) | 1 (1.3) |
| *Ethnicity* |  |  |  |  |  |
| Hispanic/Latino | 74 (9.2) | 52 (9.9) | 12 (10.1) | 6 (7.7) | 4 (5.1) |
| Not Hispanic/Latino | 728 (90.8) | 474 (90.1) | 107 (89.9) | 72 (92.3) | 75 (94.9) |
| *Comorbidities* |  |  |  |  |  |
| Heart attack | 7 (0.9) | 2 (0.4) | 1 (0.8) | 4 (5.1) | 0 (0.0) |
| Atrial fibrillation | 54 (6.7) | 28 (5.3) | 13 (10.9) | 6 (7.7) | 7 (8.9) |
| Diabetes | 107 (13.3) | 63 (12.0) | 17 (14.3) | 11 (14.1) | 16 (20.3) |
| Hypercholesterolemia | 420 (52.4) | 270 (51.3) | 67 (56.3) | 44 (56.4) | 39 (49.4) |
| Hypertension | 407 (50.7) | 253 (48.1) | 66 (55.5) | 41 (52.6) | 47 (59.5) |
| Vitamin B<sub>12</sub> deficiency | 30 (3.7) | 14 (2.7) | 8 (6.7) | 4 (5.1) | 4 (5.1) |
| Depression | 142 (17.7) | 67 (12.7) | 29 (24.4) | 21 (26.9) | 25 (31.6) |
| Anxiety | 99 (12.3) | 48 (9.1) | 22 (18.5) | 11 (14.1) | 18 (22.8) |
| Traumatic brain injury | 102 (12.7) | 63 (12.0) | 13 (10.9) | 15 (19.2) | 11 (13.9) |
| *APOE $\epsilon 4$* |  |  |  |  |  |
| One allele | 196 (24.4) | 110 (20.9) | 45 (37.8) | 21 (26.9) | 20 (25.3) |
| Two alleles | 21 (2.6) | 10 (1.9) | 3 (2.5) | 4 (5.1) | 4 (5.1) |

#### Sequence truncation and masking

Each participant’s visit history was converted into progressively truncated sequences, each containing at least two visits, and the classifiers from the main analysis were applied to every sequence. For progressors, all truncations were retained, and lead time was the conversion month (the first visit at which the diagnosis advanced) minus the month of the earliest alarm. For controls, the most recent visits were masked to reduce the chance of including participants who would progress soon after their last visit, and all truncations preceding the mask were retained. Controls left with fewer than two visits after masking had no valid sequence and were excluded.

#### Mask length

We examined the distribution of lead times of early predictions across decision thresholds and used its 80th percentile as the benchmark for mask length. For CN→MCI, about 80% of early predictions from both the non-imputed and imputed models had lead times of three visit intervals or fewer at thresholds between 0.15 and 0.50. We therefore masked three visit intervals, which excludes most near-future converters from the controls while keeping an adequate control sample; this reduced the stable CN controls from 526 to 217. For MCI→AD, about 80% of early predictions from the non-imputed models had lead times of two visit intervals or fewer at thresholds between 0.05 and 0.40, and more than 80% did so for the imputed models at every threshold. We therefore masked two visit intervals, which reduced the stable MCI controls from 78 to 23.

#### Decision thresholds

Figs S4.1 and S4.2 show sensitivity, specificity, mean lead time, and false-alarm rate as functions of the decision threshold. For CN→MCI, we selected 0.25, which keeps sensitivity high while staying close to the point below which specificity falls and the false-alarm rate rises steeply. Applying the same principle, we selected 0.30 for MCI→AD. Performance at these settings is reported in Table 5.

**Fig S4.1.**
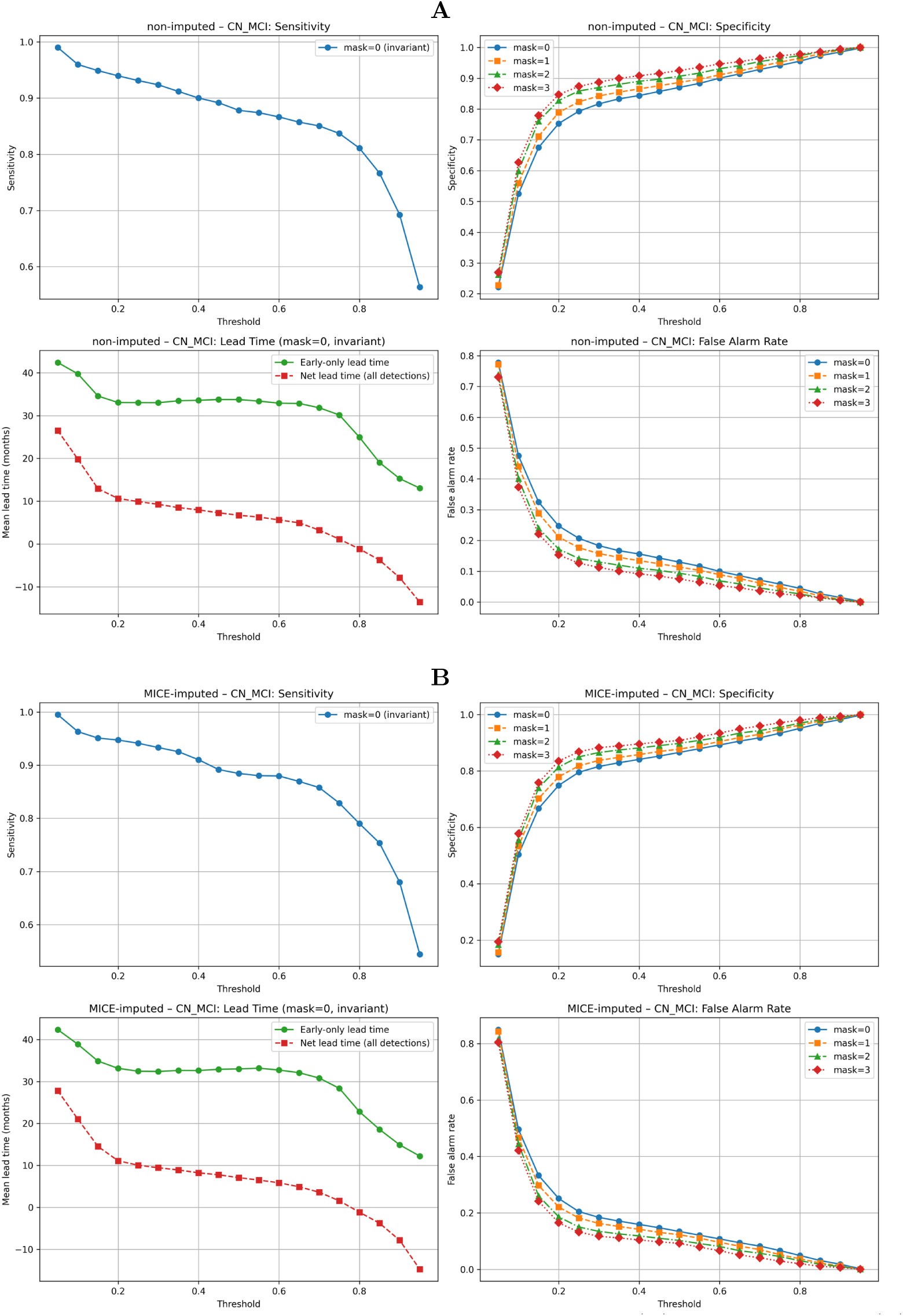
Lead-time metrics by decision threshold, CN→MCI. (A) Non-imputed and (B) MICE-imputed models. Sensitivity and lead time do not depend on the control mask; specificity and false-alarm rate are shown for mask lengths of 0–3 visit intervals. Lead time is shown for early detections only and for all detections (net).

**Fig S4.2.**
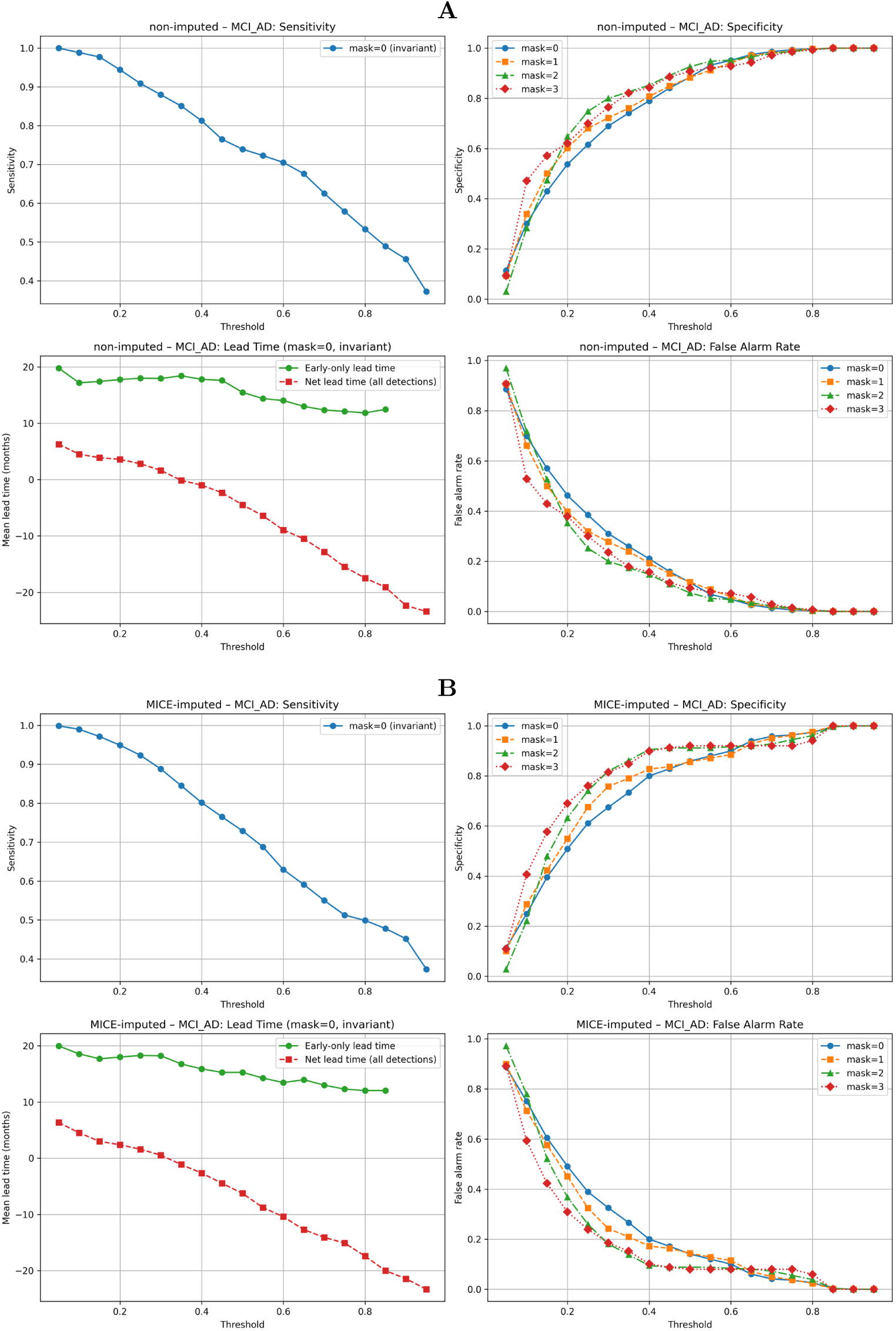
Lead-time metrics by decision threshold, MCI→AD. (A) Non-imputed and (B) MICE-imputed models. Panels as in Fig S4.1.

#### S5 Ablation study

Visit-timing differences between progressors and non-progressors, and retraining of the MCI→AD classifier without visit-timing and visit-count features.

#### Motivation

In the MCI→AD classifier, months since baseline, a visit-timing variable rather than a clinical measurement, ranked 6th of 39 variables by share of total gain (3.8%). Because Prog ID is set to 1 when any visit is more advanced than the first, and each record spans the participant’s full visit history, participants followed for longer have more opportunity to be labeled progressors. The model could therefore partly learn follow-up length rather than clinical trajectory, a form of informative censoring. We tested this by comparing visit timing between groups, by retraining the MCI→AD classifier with visit-timing and visit-count features removed, and by retraining it on each participant’s first two visits only.

#### Visit timing by progression status

In both cohorts, progressors had more visits and longer follow-up than controls (Table S5.1, Figs S5.1 and S5.2). This is consistent with how Prog ID is defined and does not by itself show that the model relies on follow-up length.

**Table S5.1.** Visit-timing statistics by progression status. Medians for controls and progressors; *p*-values from two-sided Mann-Whitney U tests.

| Variable | Control | Progressor | $p$-value |
| --- | --- | --- | --- |
| *MCI→AD* (1,396 controls, 1,437 progressors) |  |  |  |
| Visit count | 3.00 | 4.00 | $1.25 \times 10^{-75}$ |
| Follow-up span (months) | 24.18 | 42.08 | $2.21 \times 10^{-74}$ |
| *CN→MCI* (9,919 controls, 2,173 progressors) |  |  |  |
| Visit count | 4.00 | 5.00 | $6.79 \times 10^{-65}$ |
| Follow-up span (months) | 42.44 | 63.73 | $7.33 \times 10^{-75}$ |

#### Ablation design

We retrained the MCI→AD classifier under four conditions, each using the protocol of the main non-imputed analysis: the same ten stratified 80/20 train/test splits and 1,000 Optuna trials per split. Every run is therefore paired with the baseline run on the same split. Arms 1–3 removed features cumulatively after feature engineering, so all remaining features, including slopes normalized by visit time, were identical to the baseline:

- **Arm 1** removed the 11 features derived from months since baseline.
- **Arm 2** also removed the number of valid visits for each of the 19 numeric longitudinal variables (30 features removed in total).
- **Arm 3** also removed features that are missing by construction below a minimum visit count: acceleration (at least four valid visits; 19 features) and the third- and fourth-to-last observations (at least three and four valid visits; 14 features), for 63 features removed in total.

**Arm 4** kept every feature but truncated each participant’s longitudinal record to their first two visits, the cohort minimum, before feature engineering, so that all participants contributed the same number of visits. Labels were still derived from the full visit history.

**Fig S5.1.**
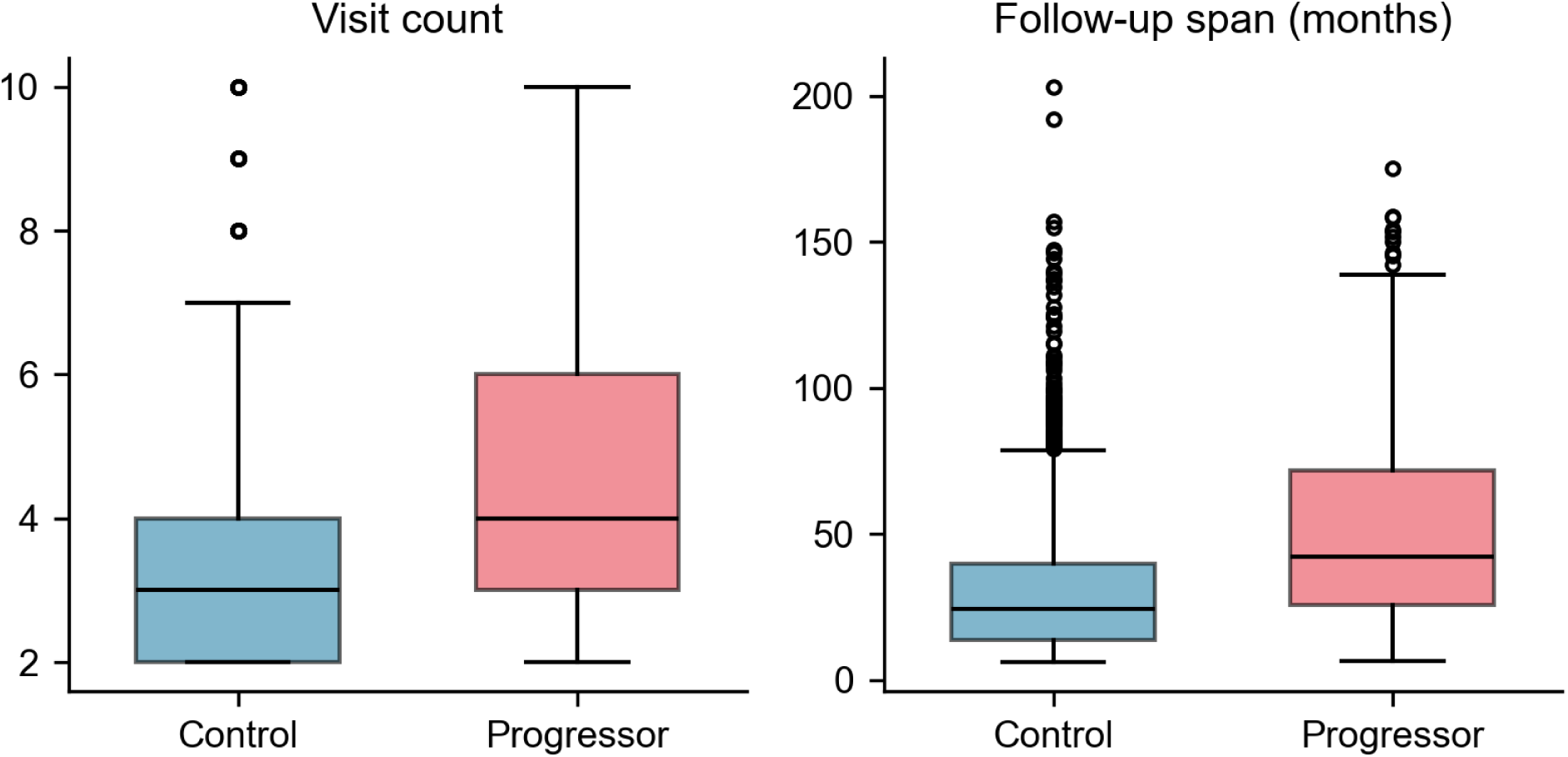
Visit timing by progression status, MCI AD cohort. Visit count and follow-up span (months) for controls and progressors.

**Fig S5.2.**
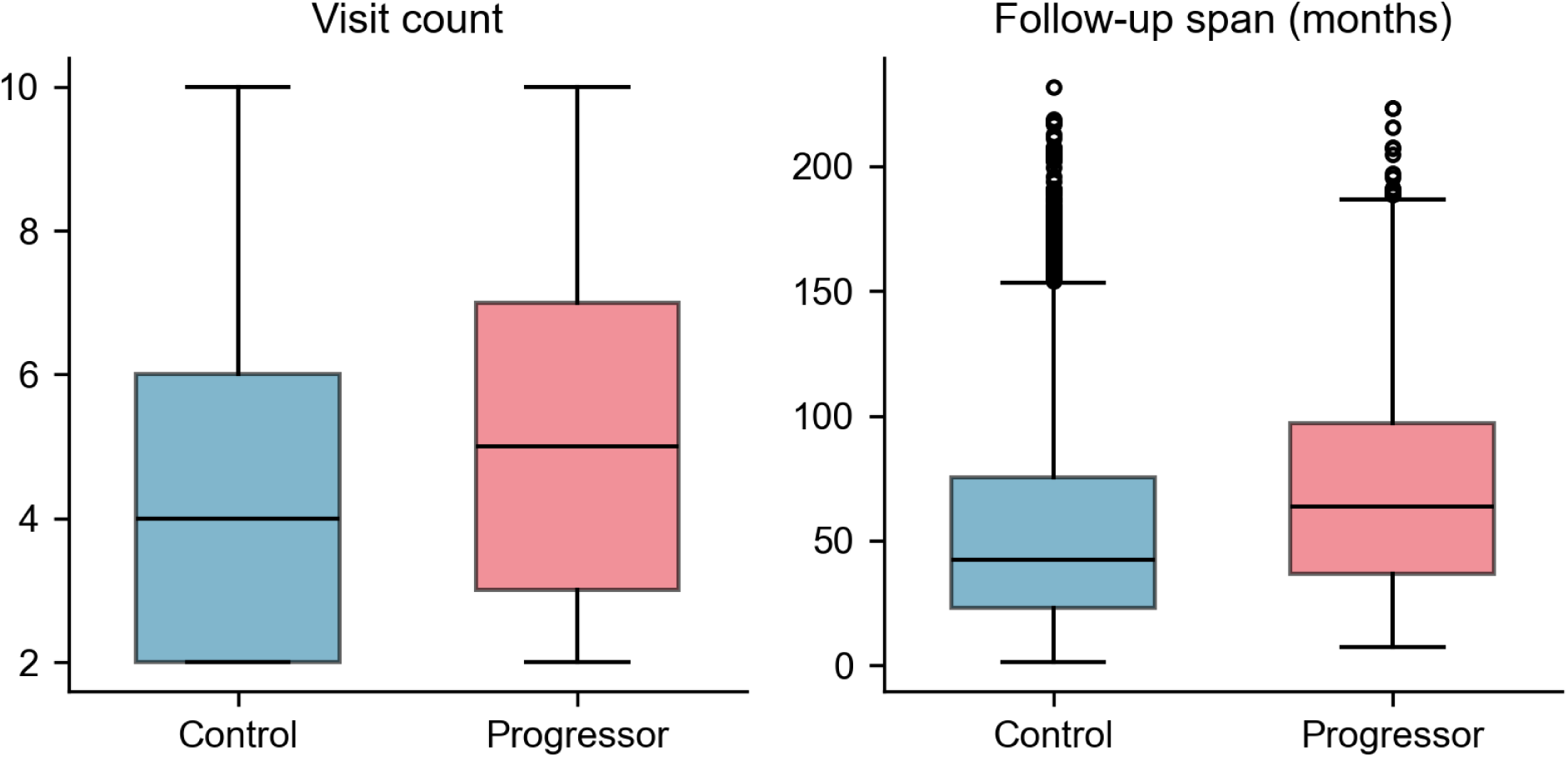
Visit timing by progression status, CN→MCI cohort. Same variables as Fig S5.1.

Because the ten splits resample the same 2,833 participants, per-split AUC differences are not independent, and standard paired tests tend to understate the variance of their mean. We therefore used the Nadeau–Bengio corrected resampled *t*-test (test-to-train ratio 0.25) [1] to obtain 95% confidence intervals for the paired difference ΔAUC = baseline − arm. For Arms 1–3 we tested equivalence against a margin of ±0.01 AUC, fixed before Arms 2–4 were trained, using two one-sided tests (TOST) [2], with Holm correction across the three arms [3]. Arm 4 removes later clinical measurements as well as differences in follow-up, so we report its AUC descriptively and make no equivalence claim. To estimate run-to-run variability of the training pipeline itself, we retrained the unmodified baseline on three of the splits.

We also audited every feature on one training split for two properties: whether its missingness alone predicted the label (AUC *>* 0.6) and its Spearman correlation with visit count. Seventeen features retained in Arm 3 had label-associated missingness: the standard deviation, slope, and slope-variability features of TOBAC30 and ALCOHOL, and the percent-change features of ten FAQ items and COMMUN. Their missingness is not determined by visit count alone. For example, TOBAC30 and ALCOHOL were not collected at UDS version 3 follow-up visits (S2), and percent change is undefined when every value before the last is zero. Arm 3 was therefore not modified; Arm 4 equalizes visit count for all participants.

## Results

In each of Arms 1–3, AUC changed by less than 0.01 (Table S5.2, Fig S5.3): the 95% confidence interval of ΔAUC included zero, and the 90% confidence interval lay within ±0.01 (Holm-adjusted TOST *p <* 0.001). The difference between the original and retrained baseline models was 0.0015, 0.0025, and −0.0002, similar in size to the differences in Arms 1–3.

Restricting all participants to their first two visits (Arm 4) reduced AUC to 0.842 ± 0.012 (ΔAUC 0.064; 95% CI 0.049 to 0.080). Across test sets, a mean of 184 of 288 progressors (64%; range 172–191) had reached AD by their second visit, so their truncated record still contains the conversion visit. After excluding these early converters from the test sets, AUC was 0.797 ± 0.022 for the truncated model and 0.929 for the baseline model on the same participants (ΔAUC 0.131; 95% CI 0.108 to 0.154).

**Table S5.2.** Discrimination by ablation arm, MCI→AD. AUC-ROC mean ± SD across ten paired splits; ΔAUC = baseline arm with Nadeau–Bengio corrected 95% CI. TOST *p*: equivalence within ±0.01, Holm-adjusted across Arms 1–3. Early converters: progressors who reached AD by their second visit.

| Model | Features | AUC-ROC | ΔAUC (95% CI) | TOST $p$ |
| --- | --- | --- | --- | --- |
| Baseline | 302 | $0.906 \pm 0.011$ | — | — |
| Arm 1: no months_since_baseline | 291 | $0.905 \pm 0.011$ | 0.0007 (−0.0018, 0.0031) | <0.0001 |
| Arm 2: + no visit counts | 272 | $0.905 \pm 0.010$ | 0.0005 (−0.0018, 0.0029) | <0.0001 |
| Arm 3: + no visit-count missingness | 239 | $0.906 \pm 0.011$ | −0.0003 (−0.0044, 0.0038) | 0.0002 |
| Arm 4: first two visits only | 302 | $0.842 \pm 0.012$ | 0.064 (0.049, 0.080) | n/a |
| excluding early converters | 302 | $0.797 \pm 0.022$ | 0.131 (0.108, 0.154) | n/a |

In Arms 1–3, CDRSUM, NACCMMSE, and COMMUN remained the three most important variables (Fig S5.4, Table S5.3). The largest single-variable gains were CDRSUM and COMMUN in Arm 1 (+0.007 and +0.006) and NACCMMSE and COMMUN in Arm 3 (+0.012 and +0.009); no variable gained more than 0.006 in Arm 2. In Arm 4, CDRSUM remained the most important variable but its share fell from 0.430 to 0.203, and COMMUN fell from 0.066 to 0.009. NACCMMSE rose from 0.099 to 0.140, and age, TOBAC30, and ALCOHOL rose to 0.102, 0.101, and 0.090. In the full-history audit, features of TOBAC30 and ALCOHOL had label-associated missingness.

**Table S5.3.** Share of total gain of selected variables by arm. Mean across the ten splits for the eight most important variables at baseline, including months since baseline, plus ALCOHOL.

| Variable | Baseline | Arm 1 | Arm 2 | Arm 3 | Arm 4 |
| --- | --- | --- | --- | --- | --- |
| CDRSUM | 0.430 | 0.437 | 0.430 | 0.423 | 0.203 |
| NACCM MSE | 0.099 | 0.101 | 0.103 | 0.111 | 0.140 |
| COMMUN | 0.066 | 0.072 | 0.068 | 0.075 | 0.009 |
| NACCBMI | 0.044 | 0.046 | 0.050 | 0.051 | 0.052 |
| TOBAC30 | 0.040 | 0.040 | 0.040 | 0.034 | 0.101 |
| months_since_baseline | 0.038 | — | — | — | 0.017 |
| NACCGDS | 0.035 | 0.037 | 0.040 | 0.039 | 0.044 |
| age | 0.035 | 0.034 | 0.035 | 0.034 | 0.102 |
| ALCOHOL | 0.024 | 0.027 | 0.019 | 0.019 | 0.090 |

### Scope of conclusions

Progressors had more visits and longer follow-up than controls, consistent with how the progression label is defined. Removing the features that directly encode visit timing and visit count (Arms 1–3) changed AUC by less than 0.01, so the reported performance does not depend on these features. This does not show that the model is free of follow-up-related signal: retained features are computed over each participant’s full history, and several correlate with visit count (e.g., CDRSUM slope variability, *ρ* = 0.74 on one training split). When visit count was equalized by truncation (Arm 4), AUC fell to 0.842, and to 0.797 when early converters were excluded. Because truncation also removes later clinical measurements, this decrease cannot be attributed to informative censoring alone.

**Fig S5.3.**
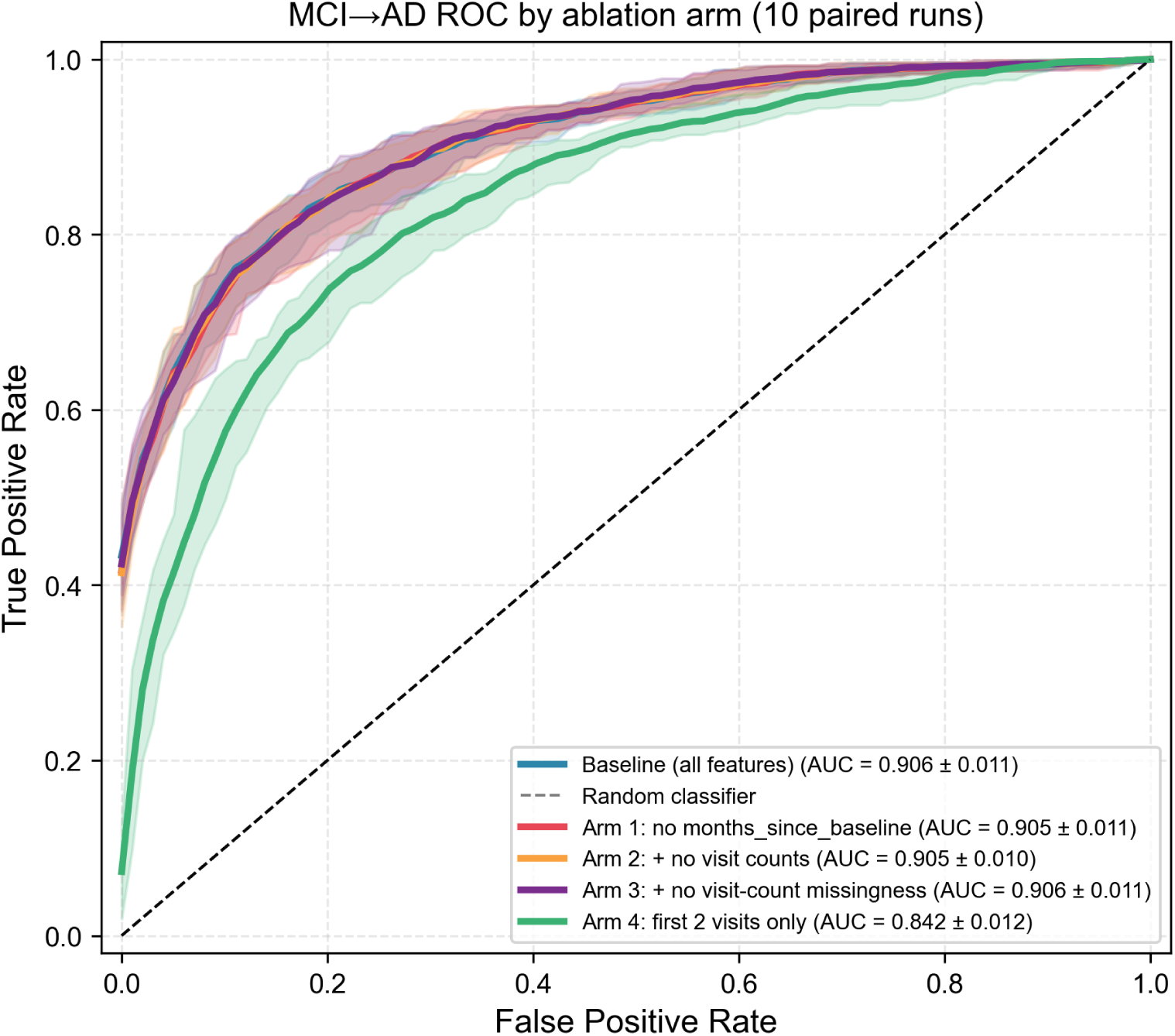
ROC curves by ablation arm, MCI AD. Mean curves across the ten paired splits; shaded bands span the 2.5th to 97.5th percentiles of the true positive rate across splits.

**Fig S5.4.**
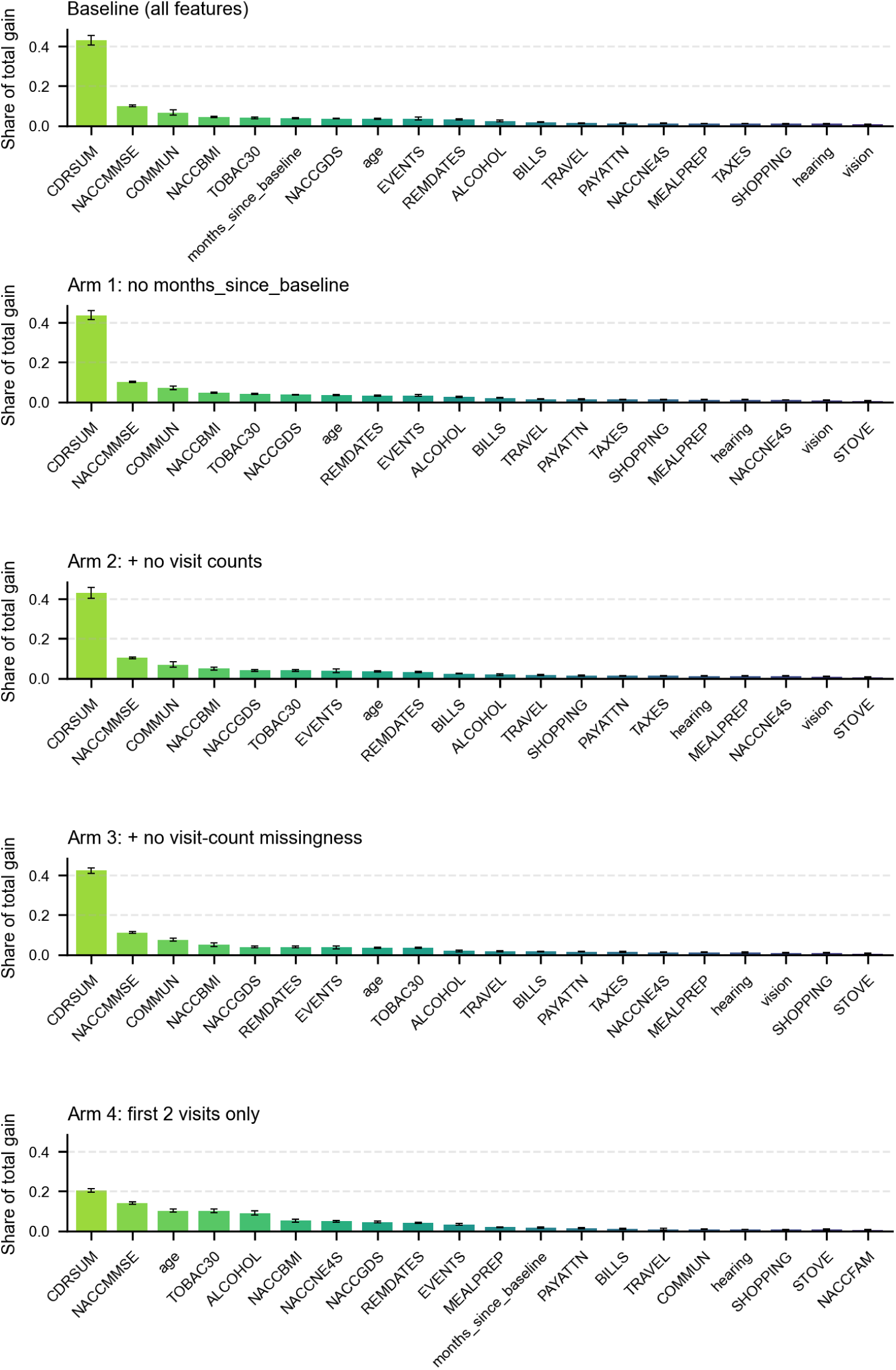
Collapsed feature importance by ablation arm, MCI→AD. Top 20 variables per arm, each variable’s share of total gain summed across all features derived from it (e.g., CDRSUM slope and CDRSUM last). Bars show the mean share across the ten splits; error bars show a 95% percentile bootstrap confidence interval of the mean.

## S6 Additional classification results

ROC and precision–recall curves of non-imputed and MICE-imputed models, feature importance and collapsed SHAP summaries of the MICE-imputed models, and non-imputed results for individual derived features.

## Non-imputed and MICE-imputed models

The ROC and precision–recall curves of the non-imputed and MICE-imputed models overlap for both tasks (Fig S6.1), consistent with the AUC-ROC and PR-AUC values in Table 4.

Feature rankings of the MICE-imputed models closely matched those of the non-imputed models, with a Spearman correlation of the variables’ shares of total gain of 0.97 for CN→MCI and 0.92 for MCI→AD (Fig S6.2). CDRSUM and NACCMMSE again ranked first and second in both tasks, both by total gain and by mean absolute SHAP value (Figs S6.3 and S6.4). Table S6.1 gives the share of the SHAP attribution of selected variables and of each feature type for both pipelines.

## Collapsed SHAP plots

The collapsed SHAP plots in Figs 4–5 and S6.3–S6.4 summarize attribution at the underlying-variable level. For each training observation, SHAP values for engineered features derived from the same variable were summed, and the resulting observations were pooled across the ten runs for each task and pipeline. Each dot represents one observation–model prediction; its horizontal position shows the variable’s signed contribution to the predicted log-odds. For variables with multiple derived features, color encodes an approximate group-level value: constituent feature values were min–max normalized and combined using absolute SHAP values as weights, with an unweighted mean used when all weights were zero. Rows follow a shared variable order across the collapsed plots, and vertical offsets show the density of observations with similar SHAP values.

## Individual derived features

Figs S6.5–S6.7 show the 50 individual engineered features of the non-imputed models with the highest share of total gain and mean absolute SHAP value, before summing over each variable’s derived features as in Figs 3–5.

**Fig S6.1.**
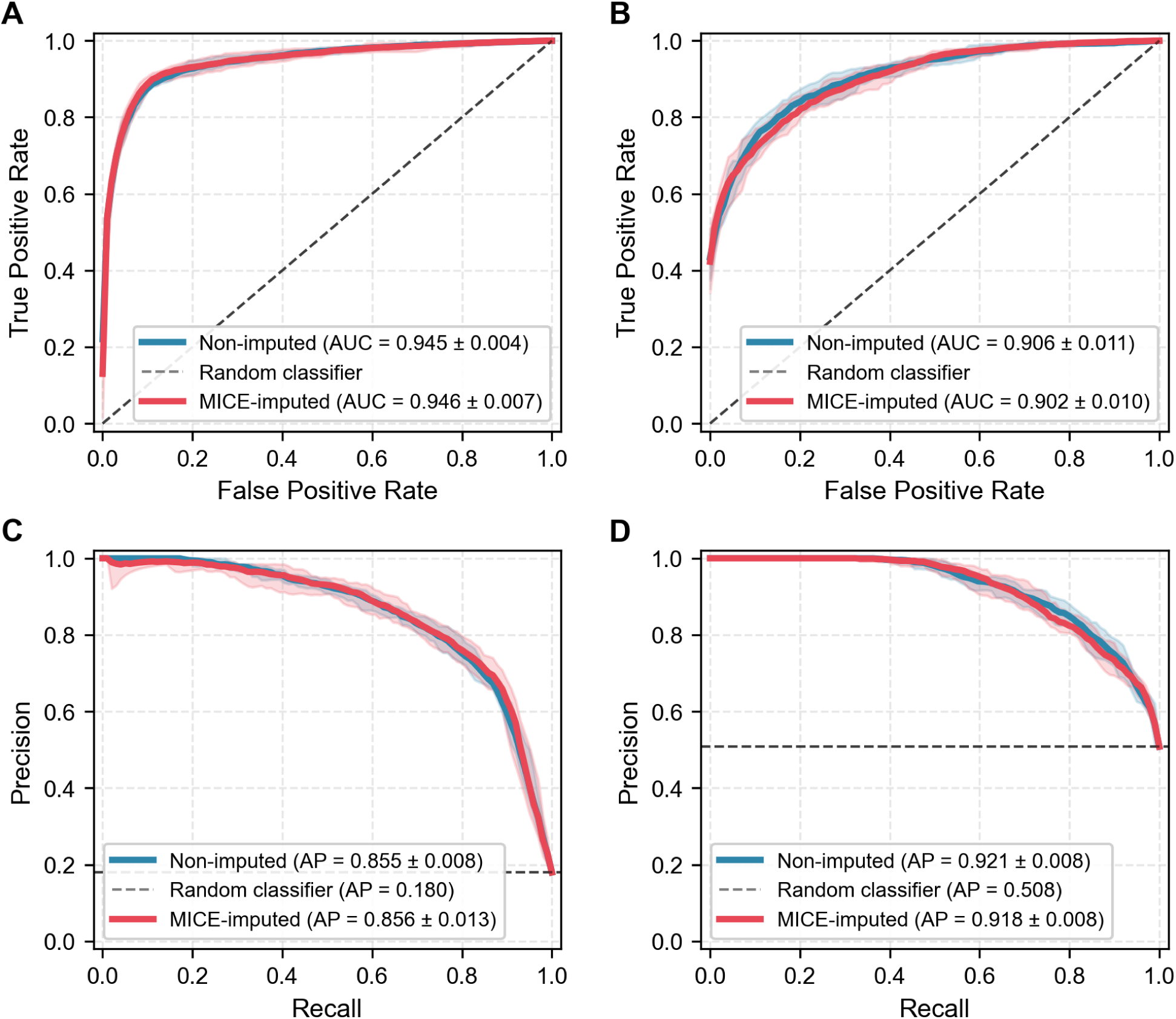
ROC and precision–recall curves of non-imputed and MICE-imputed models. (A, B) ROC and (C, D) precision–recall curves for CN→MCI (A, C) and MCI→AD (B, D). Solid lines show the mean curve across the ten runs of each pipeline; shaded bands span the 2.5th to 97.5th percentiles of the per-run curves. Legend values are mean ± SD across runs. Dashed lines show a random classifier, whose average precision equals the positive-class prevalence.

**Table S6.1.**
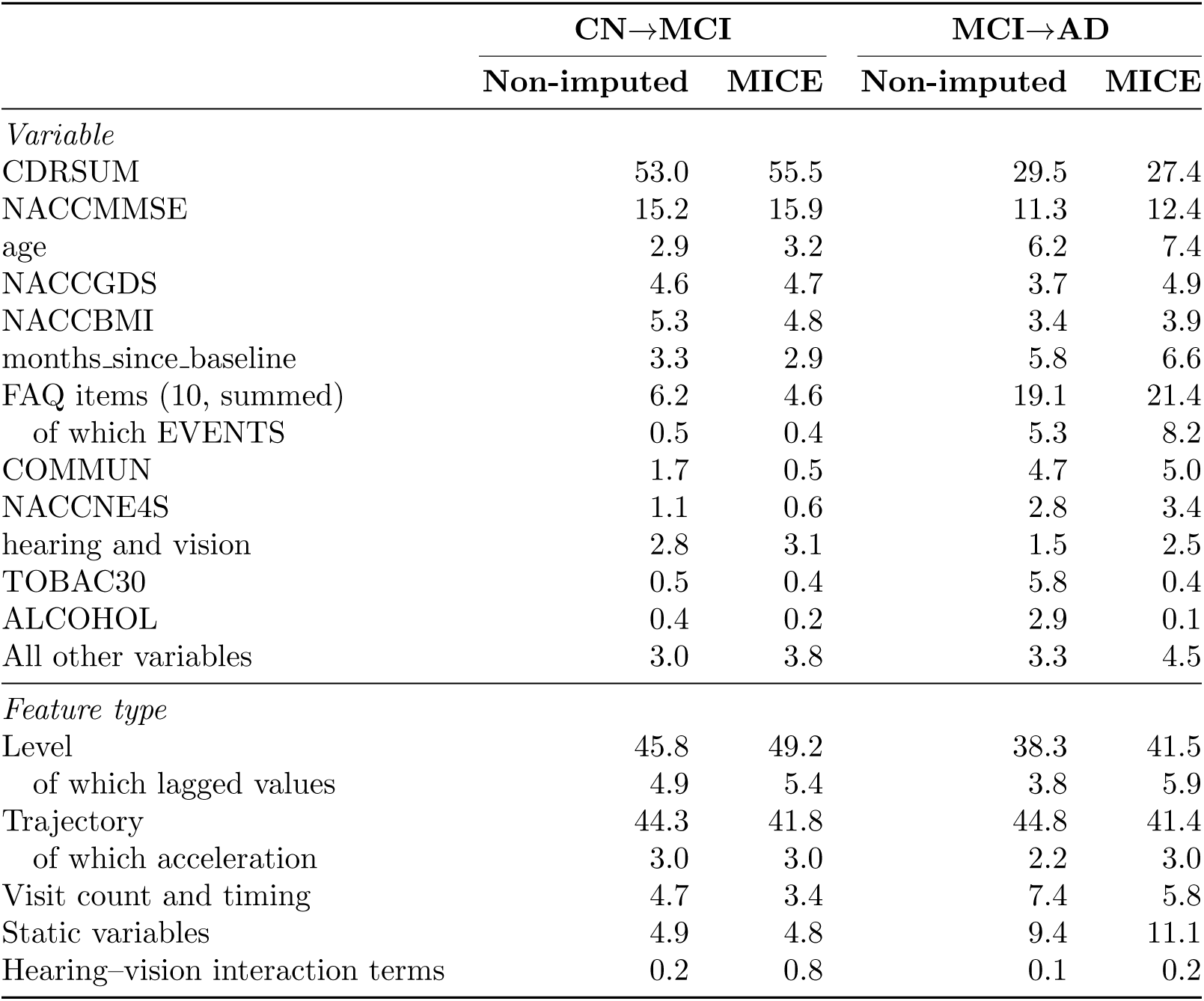
Share of SHAP attribution by variable and feature type (%). Computed on training samples pooled across the ten runs of each pipeline. Variables: mean absolute SHAP value after summing SHAP values over the variable’s derived features, as a percentage of the total over all 39 variables. Feature types: mean absolute SHAP values of individual features summed by type, as a percentage of the total over all 302 features. Level features are the first, last, mean, maximum, minimum, and lagged values and the living-situation change flag; trajectory features are the slope, acceleration, last minus first, range, standard deviation, slope variability, and percentage change; visit count and timing comprises the numbers of valid visits and the features derived from months since baseline.

**Fig S6.2.**
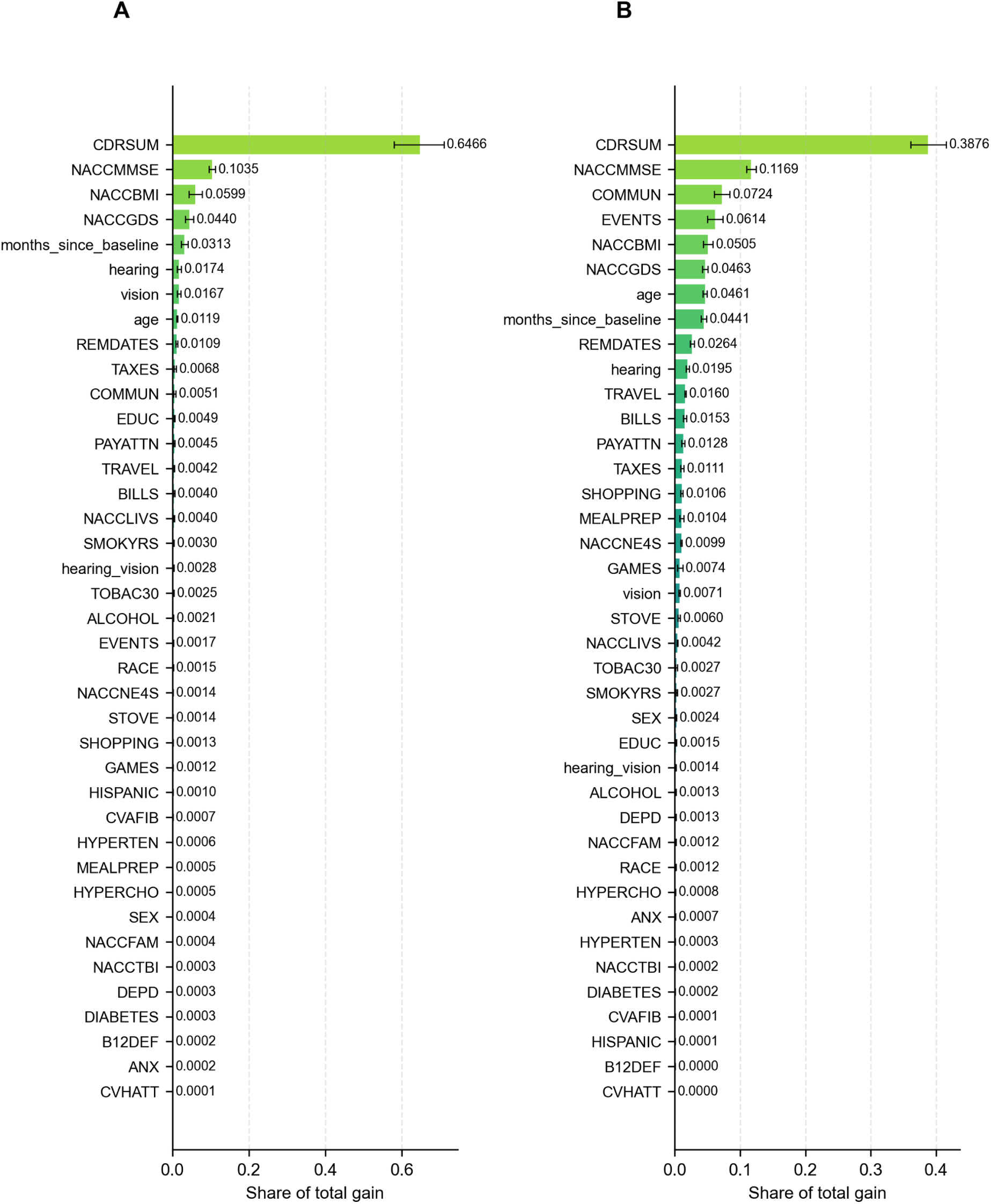
Feature importance of the MICE-imputed models. All 39 variables ranked by their share of the total gain, averaged across the ten MICE-imputed runs, for (A) CN→MCI and (B) MCI→AD classification. A variable’s total gain is the loss reduction from all splits on the features derived from it. Bars show the mean share across runs and error bars denote the 95% percentile bootstrap confidence interval of the mean (2,000 resamples of the ten runs).

**Fig S6.3.**
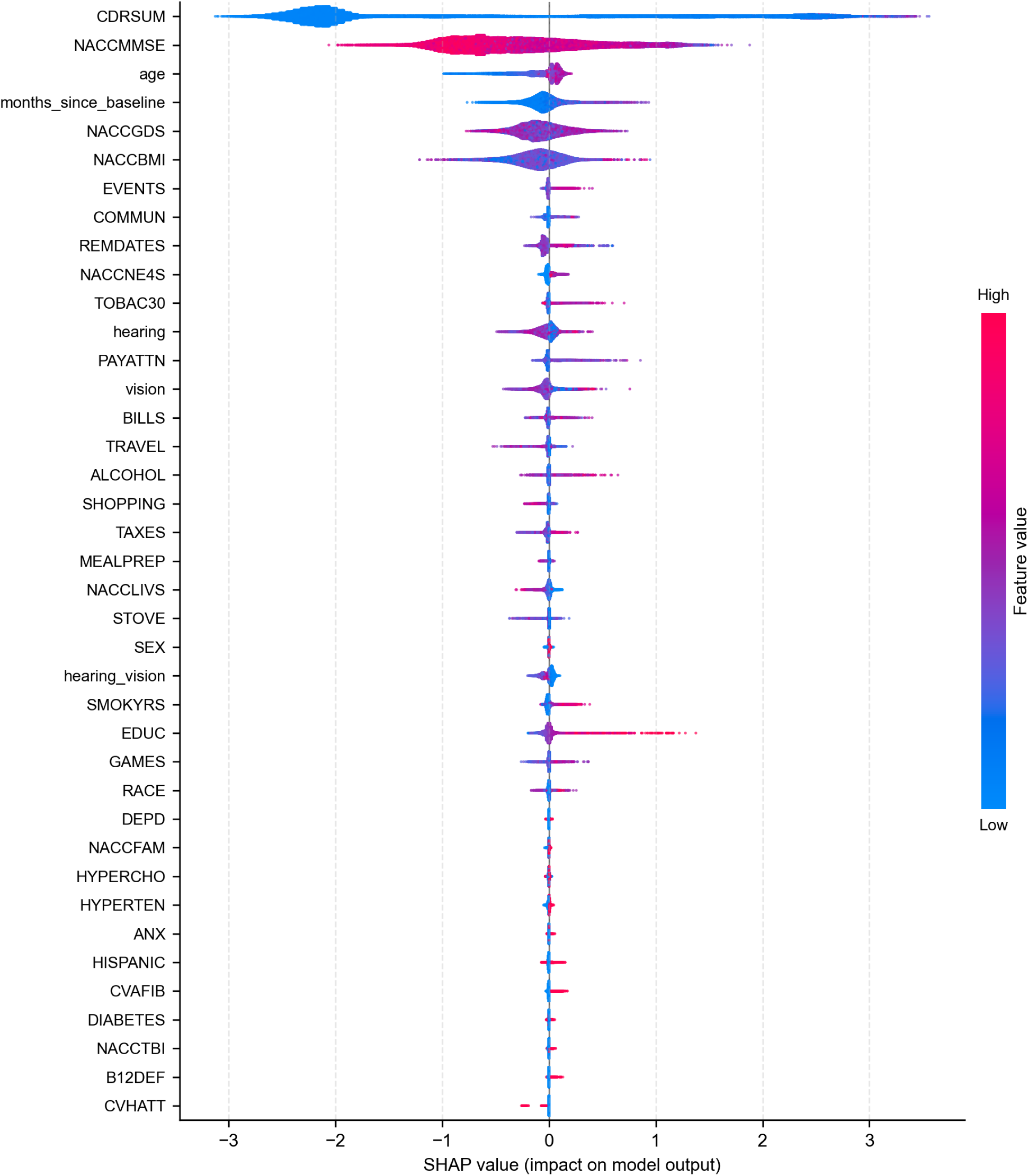
SHAP summary plot for the MICE-imputed CN→MCI classifier. All 39 variables, with SHAP values computed on training samples pooled across the ten MICE-imputed runs and summed across all features derived from each variable. Dots, colors, and row order as in Fig 4.

**Fig S6.4.**
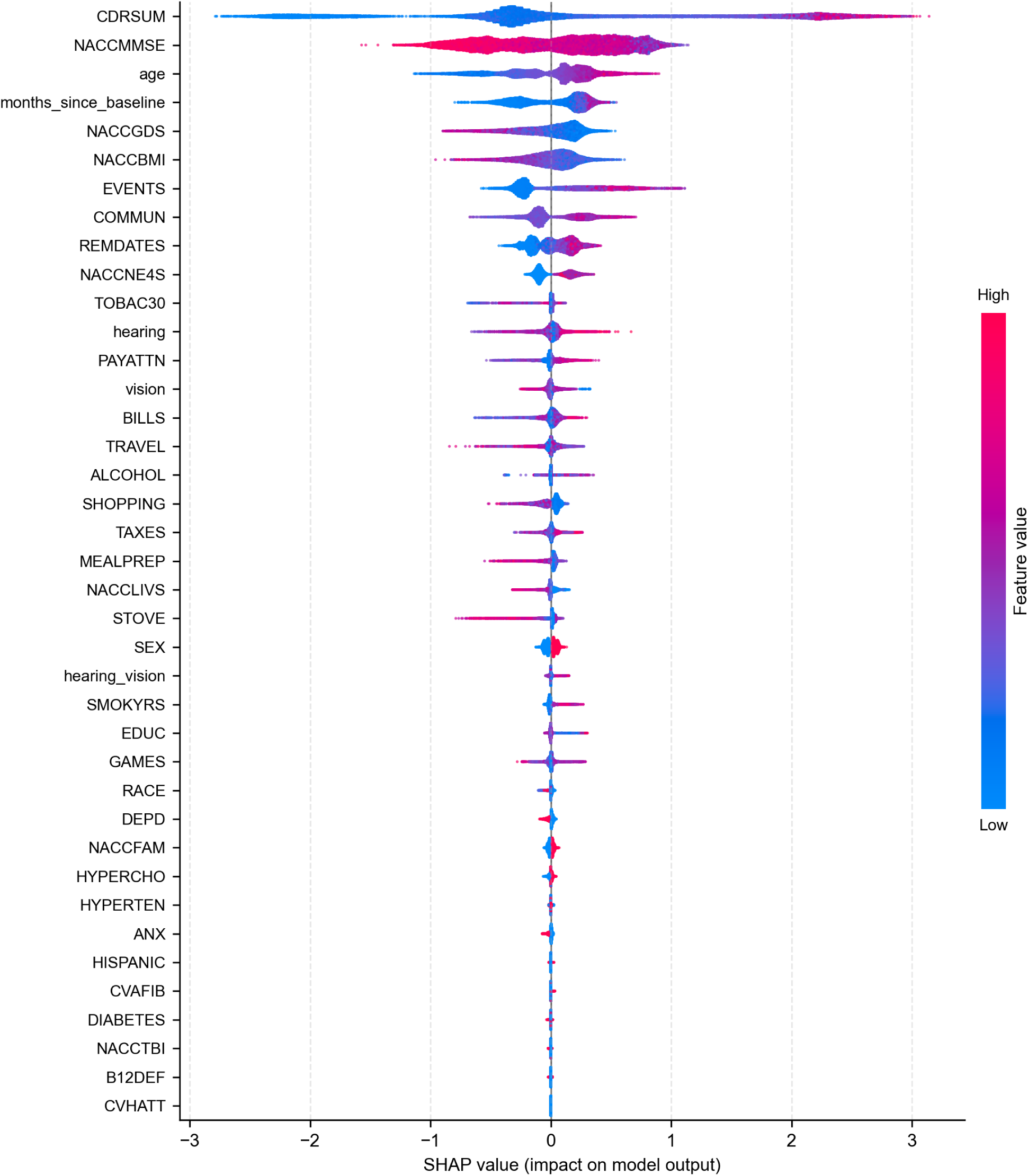
SHAP summary plot for the MICE-imputed MCI→AD classifier. All 39 variables, with SHAP values computed on training samples pooled across the ten MICE-imputed runs and summed across all features derived from each variable. Dots, colors, and row order as in Fig 4.

**Fig S6.5.**
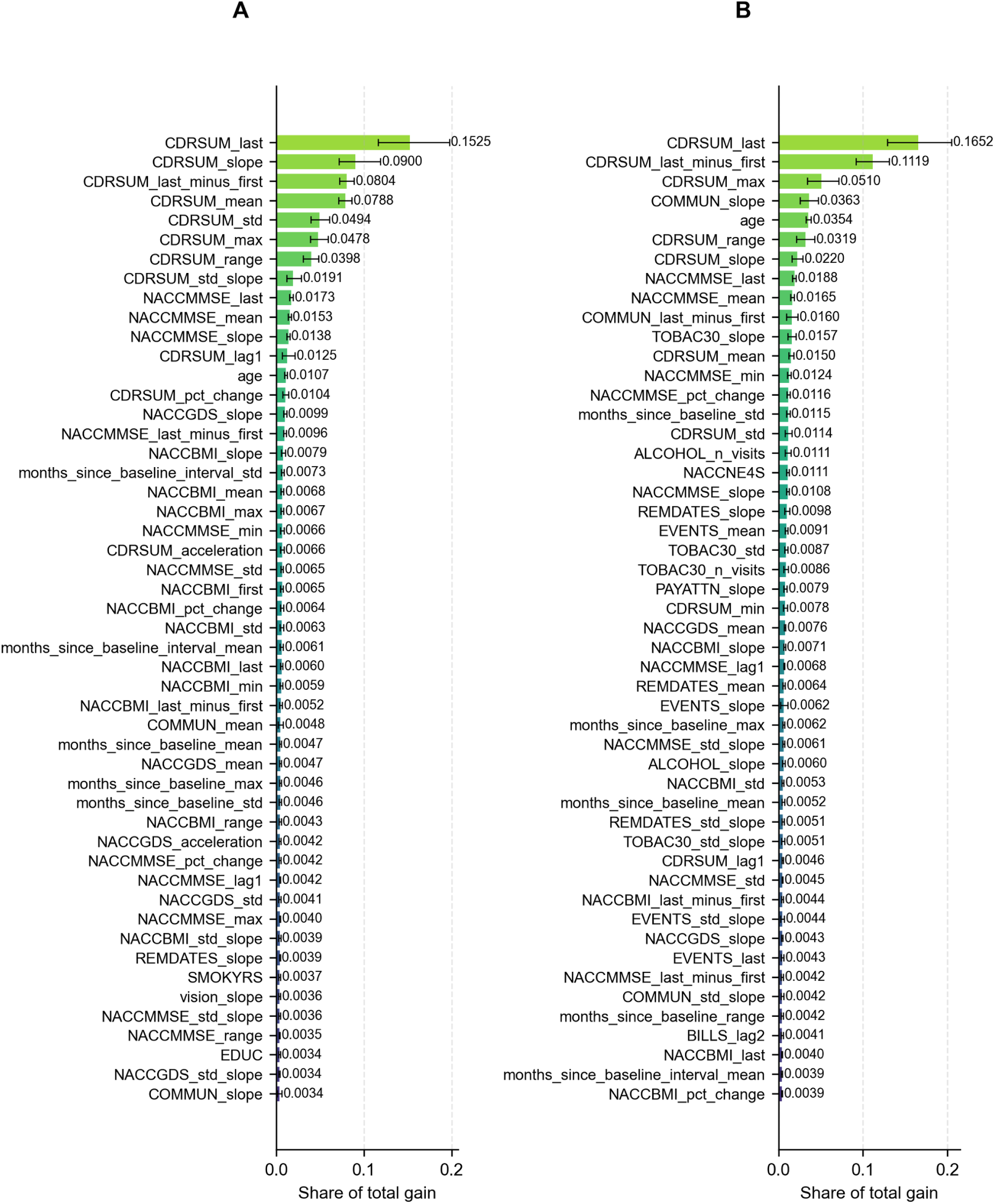
Feature importance of individual derived features. Top 50 features ranked by their share of the total gain, averaged across the ten non-imputed runs, for (A) CN→MCI and (B) MCI→AD classification. Bars show the mean share across runs and error bars denote the 95% percentile bootstrap confidence interval of the mean (2,000 resamples of the ten runs).

**Fig S6.6.**
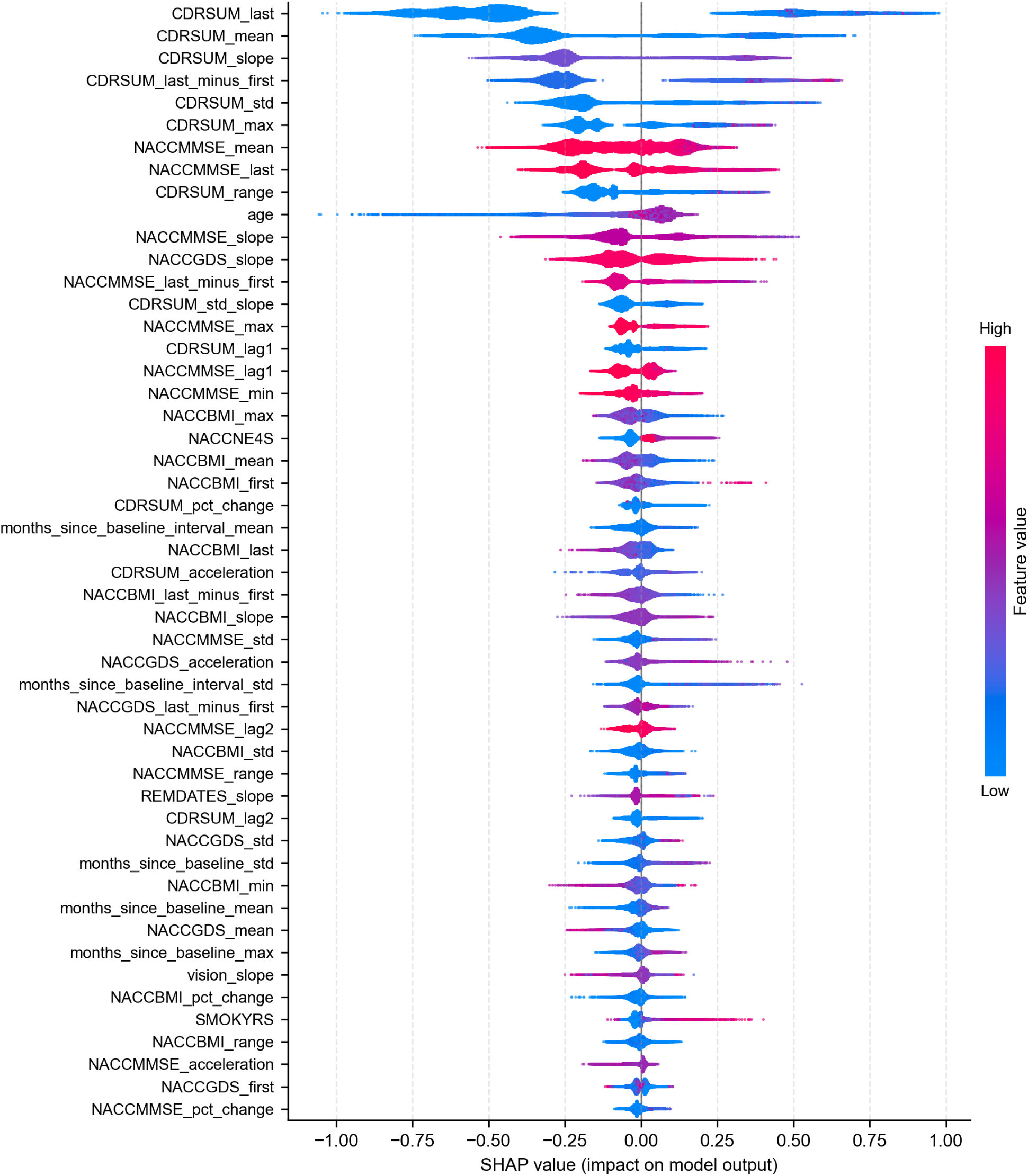
SHAP summary plot of individual derived features, CN→MCI. The top 50 features are ranked by mean absolute SHAP value computed on training samples pooled across the ten non-imputed runs. Each dot represents a single training prediction; horizontal position indicates the feature’s contribution to the predicted log-odds, and dot color indicates the feature value (low to high).

**Fig S6.7.**
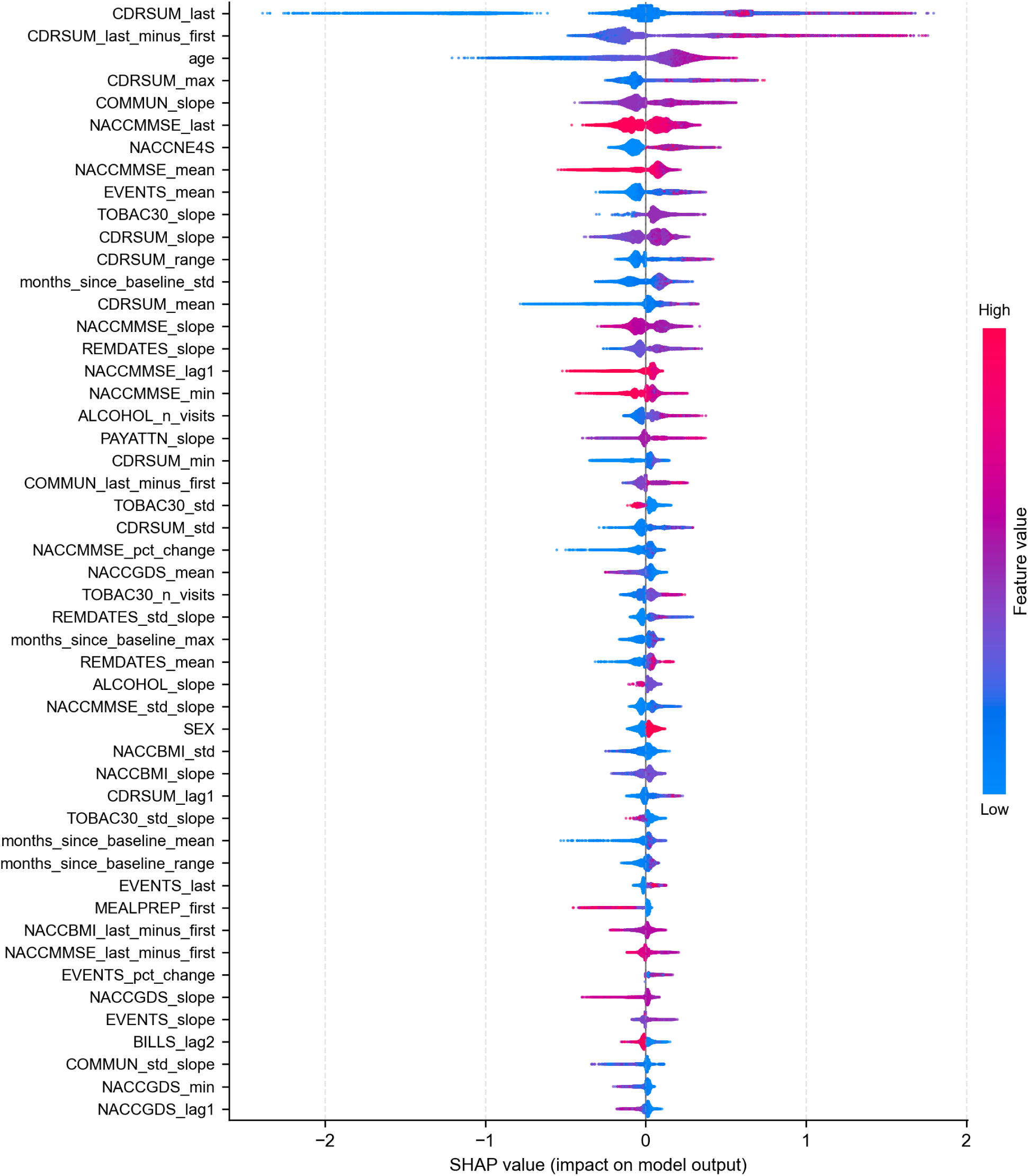
SHAP summary plot of individual derived features, MCI→AD. The top 50 features are ranked by mean absolute SHAP value computed on training samples pooled across the ten non-imputed runs. Each dot represents a single training prediction; horizontal position indicates the feature’s contribution to the predicted log-odds, and dot color indicates the feature value (low to high).

## Notes

### Competing Interest Statement

The authors have declared no competing interest.

